# Covid-19 Exposure Assessment Tool (CEAT): Easy-to-use tool to quantify exposure based on airflow, group behavior, and infection prevalence in the community

**DOI:** 10.1101/2022.03.02.22271806

**Authors:** Brian J. Schimmoller, Nídia S. Trovão, Molly Isbell, Chirag Goel, Benjamin F. Heck, Tenley C. Archer, Klint D. Cardinal, Neil B. Naik, Som Dutta, Ahleah Rohr Daniel, Afshin Beheshti

**Affiliations:** Signature Science LLC, Austin, TX, 78759, USA; COVID-19 International Research Team; Fogarty International Center, National Institutes of Health, Bethesda, Maryland, USA; Northwestern University Feinberg School of Medicine, Chicago, IL, 60611, USA; Bastion Technologies, NASA Ames Research Center, Moffett Field, CA, 94035, USA; Biomea Fusion, Inc. Redwood City, CA, 94063, USA; Leidos, Inc., NASA Ames Research Center, Moffett Field, CA, 94035, USA; Mechanical & Aerospace Engineering, Utah State University, Logan, UT 84332, USA; Space Biosciences Division, NASA Ames Research Center, Moffett Field, CA, 94035, USA; KBR, Space Biosciences Division, NASA Ames Research Center, Moffett Field, CA, 94035, USA; Stanley Center for Psychiatric Research, Broad Institute of MIT and Harvard, Cambridge, MA, 02142, USA

**Keywords:** COVID-19, SARS-CoV-2, aerosol model, COVID-19 Exposure Assessment, CEAT, Wells-Riley, Eddy Diffusivity

## Abstract

The COVID-19 Exposure Assessment Tool (CEAT) allows users to compare respiratory relative risk to SARS-CoV-2 for various scenarios, providing understanding of how combinations of protective measures affect exposure, dose, and risk. CEAT incorporates mechanistic, stochastic and epidemiological factors including the: 1) emission rate of virus, 2) viral aerosol degradation and removal, 3) duration of activity/exposure, 4) inhalation rates, 5) ventilation rates (indoors/outdoors), 6) volume of indoor space, 7) filtration, 8) mask use and effectiveness, 9) distance between people, 10) group size, 11) current infection rates by variant, 12) prevalence of infection and immunity in the community, 13) vaccination rates of the community, and 14) implementation of COVID-19 testing procedures. Demonstration of CEAT, from published studies of COVID-19 transmission events, shows the model accurately predicts transmission. We also show how health and safety professionals at NASA Ames Research Center used CEAT to manage potential risks posed by SARS-CoV-2 exposures. Given its accuracy and flexibility, the wide use of CEAT will have a long lasting beneficial impact in managing both the current COVID-19 pandemic as well as a variety of other scenarios.

## Introduction

The novel severe acute respiratory syndrome coronavirus 2 (SARS-CoV-2) that causes the coronavirus disease 2019 (COVID-19) has quickly spread around the world and was formally recognized as a pandemic by the World Health Organization (WHO) on March 11, 2020 (WHO, 2021 March 11). COVID-19 poses a great public health, clinical, economical, and societal burden worldwide. SARS-CoV-2 transmission occurs mainly through close contact (WHO, 2021 March 11), by direct and indirect contact (via fomites), and through the air via respiratory droplets and/or airborne particles (i.e., aerosols) [1].

The global and local transmission dynamics drove an urgent need for assessing potential risk of transmission while performing different activities in various facilities. Public health officials have had to reevaluate how the public should interact to reduce and contain viral spread [2–5], leading to assessment of worker and group risks associated with viral exposure in various settings [6–9]. Risk assessment and planning regularly consider the contribution of an array of factors, using largely qualitative guidance from public health and media sources [3,9,10], such as the viral exposure pathways, risk of infection (e.g., number of cases per population), efficacy of interventions and personal protective equipment (PPE **–** e.g., masks, gloves), human behavior (e.g., adhering to public health guidelines, hand washing, social distancing), and environmental factors (e.g., ventilation). Given the numerous factors that affect exposure to the virus, qualitative assessments are insufficient when trying to compare various courses of action or potential mitigation options.

The WHO and the Centers for Disease Control and Prevention (CDC) have released guidance for risk assessment and management of exposure in the context of COVID-19 at work [11], towards health-care personnel [12], community-related [13], and associated with domestic and international travel [14]. However, the qualitative nature of these guidelines makes it difficult to quantify the exposure risk in varied settings.

Researchers have developed tools that predict the risk of transmission from exposure through inhalation of emitted SARS-CoV-2-containing aerosols. Risk assessment tools provide an important means of gaining understanding of dynamics of transmission and evaluating and comparing risks associated with local environmental conditions, community epidemiological factors, and mitigation options. Typically, the infectious disease risk assessment tools utilize either a deterministic dose-response approach or, alternatively, a Wells-Riley approach [15]. A detailed comparison of dose-response models and Wells-Riley models applied to infectious disease risk assessment is provided in Sze and Chao, 2010, addressing both models’ advantages and limitations. Specific to SARS-CoV-2, Miller et al. (2021) [16] offers a Wells-Riley-based method to model transmission and has developed a companion COVID-19 Aerosol Transmission Estimator spreadsheet-based tool [17]. The Wells-Riley-based method addresses physical factors that contribute to indoor transmission, applying a uniform well-mixed box (WMB) assumption and transmission estimates using the Wells-Riley equation. Bazant and Bush, 2021 [18] provide a comprehensive physical model of the factors that affect indoor transmission and released a spreadsheet-based tool and online app [19] that calculates safety guidelines to limit the viral transmission based upon Wells-Riley and WMB assumptions. This tool recommends the total number of hours of exposure that are permissible given the number of people, their behavior, characteristics of the room and its ventilation, and the prevalence of COVID-19 and variants in the community. Parhizkar, et al., 2021 [20] developed a dose-response approach and model that uses a WMB assumption and a novel treatment of the inhaled and deposited doses. They demonstrate the model’s capability against well-documented COVID-19 outbreaks and offer a demo version of an online tool [21]. Wagner, et al., 2021 [22] offer a comprehensive modeling study that examines both indoor and outdoor exposures from two-person interactions, examining near-field and far-field effects, and modeling the behavior of particulates of various sizes. Other modeling efforts have focused on predicting transmission risks using epidemiological and behavioral factors and population statistics [23–25], without addressing facility- and event-specific physical mechanisms that would affect transmission risk.

Rigorous study of the physics of aerosol behavior in indoor spaces has also been accomplished using both experiments and computational fluid dynamic (CFD) numerical simulations. These studies analyzed key aspects of the hydrodynamics produced by expiratory events, including sneezing, coughing, talking, singing and breathing, and the dispersion processes of the resulting aerosol cloud [26–28]. While these experiments and modeling studies produce important understanding of the aerosol behavior in the environment and the respiratory system, their results are specific to the defined scenarios that were modeled and are too computationally intensive to be used directly and routinely by non-experts.

Our goal was to develop a simple-to-use, quantitative exposure and risk assessment tool that addresses the factors summarized **Fig. 1** and was based on principles of epidemiological, physics, and engineering to provide benefit to risk assessors and decision makers in a variety of settings. Additionally, we wanted to incorporate recent findings regarding disease characteristics and virus dynamics Our focus was to create a tool that could be easily used by people who are tasked with making recommendations or decisions for their organizations or groups (e.g., businesses, schools, and civic groups) on approaches to reduce viral exposure. The end result of our project has been the development of the COVID-19 Exposure Assessment Tool (CEAT). The CEAT model is embedded in an Adobe® PDF (Portable Document Format) file and was coded in JavaScript using Adobe Acrobat’s ® “Prepare a Form” function. The model’s user interface is shown in **Fig. 2A** and is available for download at https://www.cov-irt.org/exposure-assessment-tool/ as a PDF. The PDF platform was chosen instead of a web app, since the PDF allows organizations to use the tool within the privacy and security of their own networks and devices, eliminating any concern that an organization’s private worker safety information was being shared externally. Additionally, the PDF offers the ability to save and disseminate the results for specific events and scenarios as individual PDF files. The underlying algorithm used in CEAT leverages aspects of both Wells-Riley models and dose-response models. An important difference between the CEAT model and the other models discussed above is that CEAT assesses the additional higher concentration of virus containing aerosols that may occur when people are in close proximity and applies this approach to groups between 2 and 250 people, both indoors and outdoors. The model relies on information that the users would have available or could reasonably estimate, addresses the mechanisms that are within the organization’s control (e.g., distancing, duration, ventilation rates, filtration, mask wearing, vaccination requirements, and option for indoor/outdoor activities), and communicates a clear and easily interpretable result. The model attempts to address the full range of exposure risks within a community, from highest-exposure risk to people known to be infected, typical of a clinical environment, to lowest-risk exposure to people who rigorously follow public health guidance.

**Figure 1.**
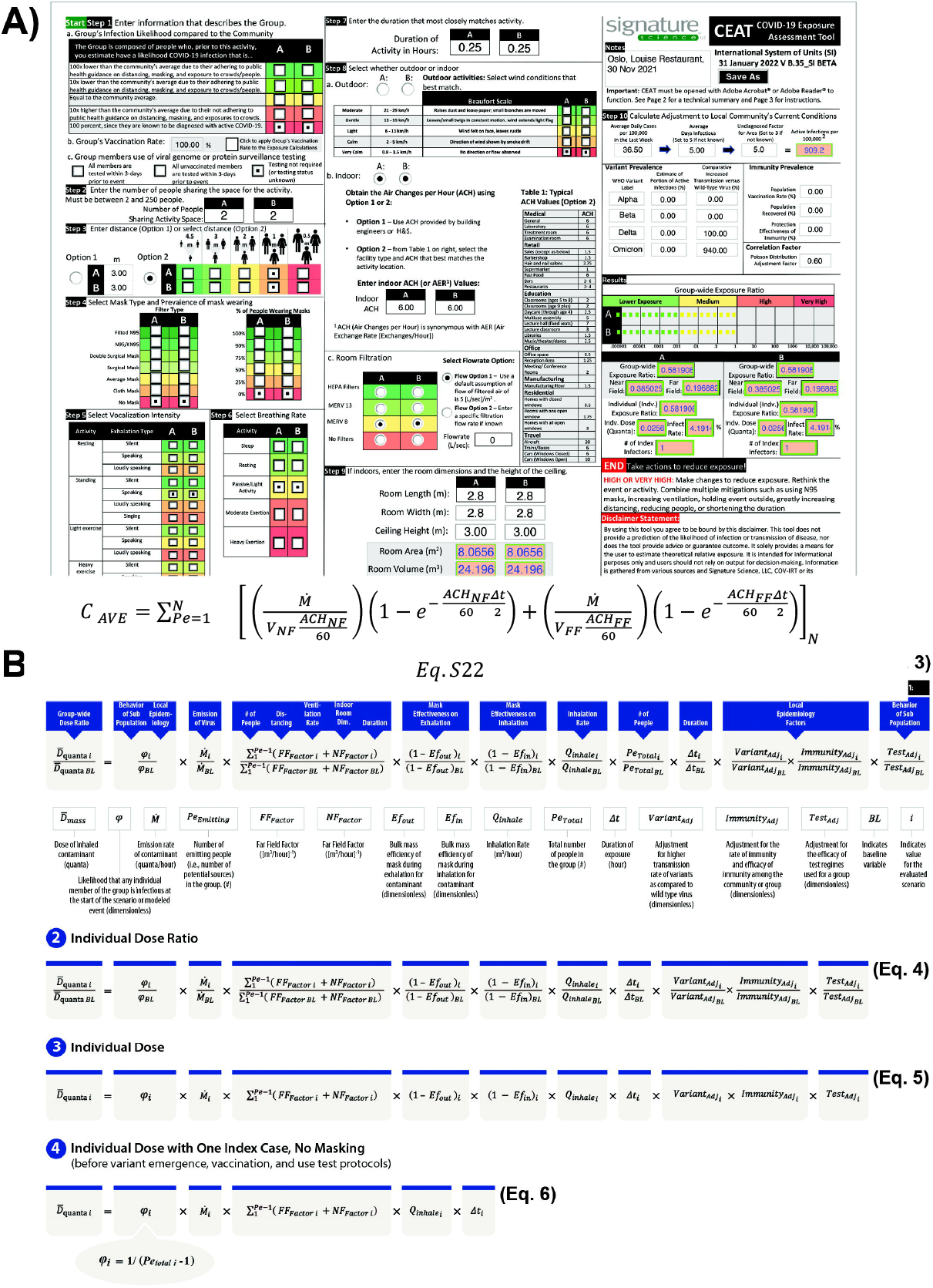
COVID-19 Exposure Assessment Tool Interface (CEAT) and background on the model utilized. **A)** User interface of the interactive PDF for CEAT. **B)** The equations that the CEAT model uses to calculate results.

**Figure 2.**
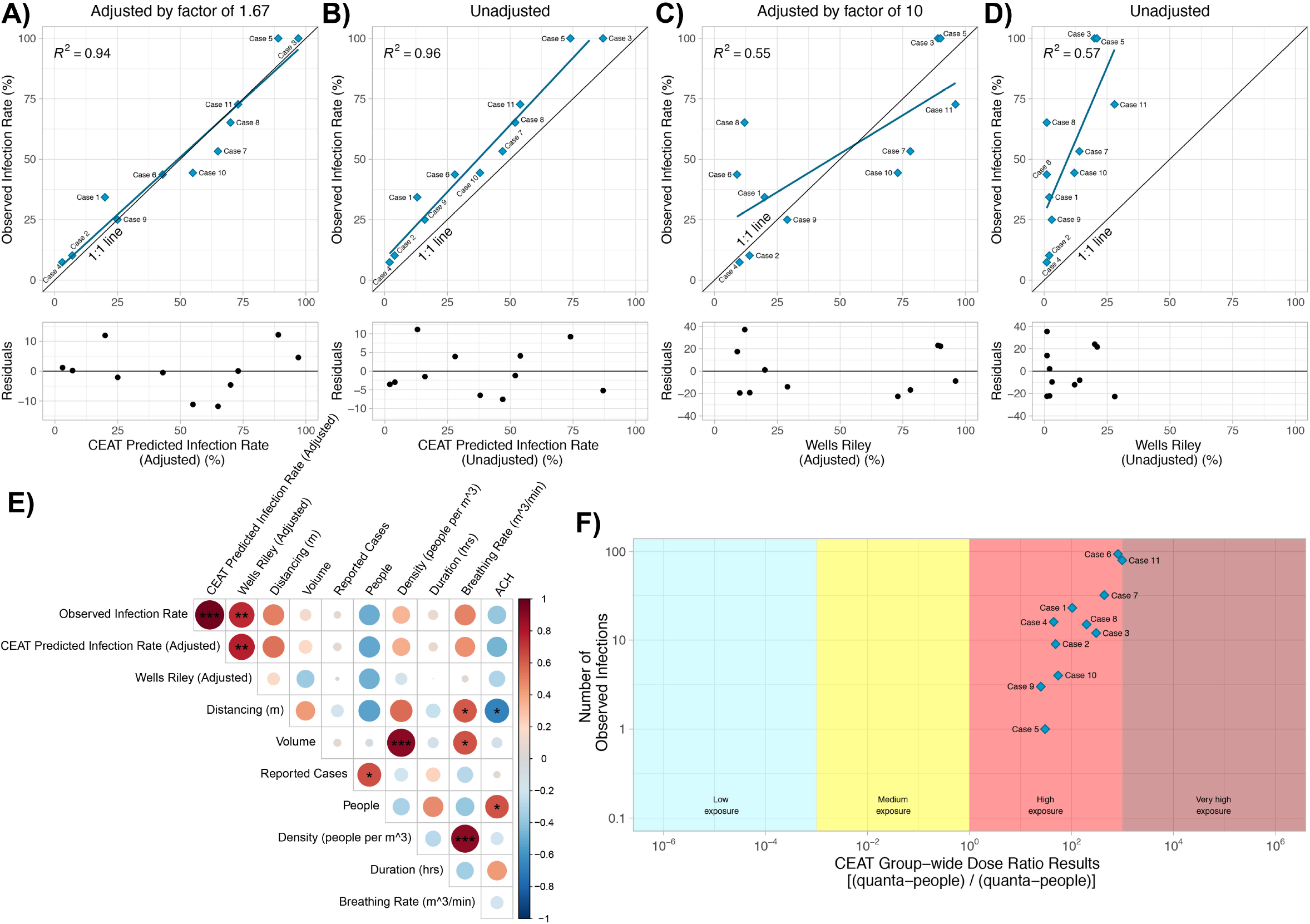
Validation of the CEAT with Known COVID-19 Spreading Events. **A)** and **B)** The adjusted and unadjusted scatter plot comparing the observed infection rates of known events (found in **Table 2**) to CEAT predicted infection rates. **C)** and **D)** The adjusted and unadjusted scatter plot comparing the observed infection rates of known events to Wells-Riley model predicted infection rates. For **A) - D)** linear fits were made to the data points and the residuals of these fits are plotted underneath each plot. The R^2^ values for the fits are shown in the plots. **E)** Correlation plot of the observed infection rate to both the CEAT and Wells-Riley adjusted predicted infection rates. Correlation with additional parameters from the event is shown. The size of the nodes reflects the degree of correlation (i.e., larger the size the higher the correlation). Positive correlation is related to the higher shades of red, while negative correlation is related to higher shades of blue. Statistically significant correlations are denoted by *** p-value < 0.001, ** p-value < 0.01, and * p-value < 0.05. **F)** Scatter plot of the exposure risk for all eleven events determined by CEAT.

## Results

### Model Overview

CEAT allows users to estimate group-wide and individual relative dose, an individual dose, and transmission risk from potential SARS-CoV-2 exposure in various scenarios, based on the key mechanistic, viral, and epidemiological factors summarized in **Fig. S1A** and **Table 1**. Here we present 1) a brief overview of the CEAT model; 2) the demonstration of the model applied to real-world, well-documented transmission scenarios; 3) describe how CEAT was applied operationally by NASA Ames Research Center’s Health and Safety office to manage exposure risk of its staff. Full details of the mathematical model used for CEAT can be found in the **STARS Methods** section.

**Table 1.**
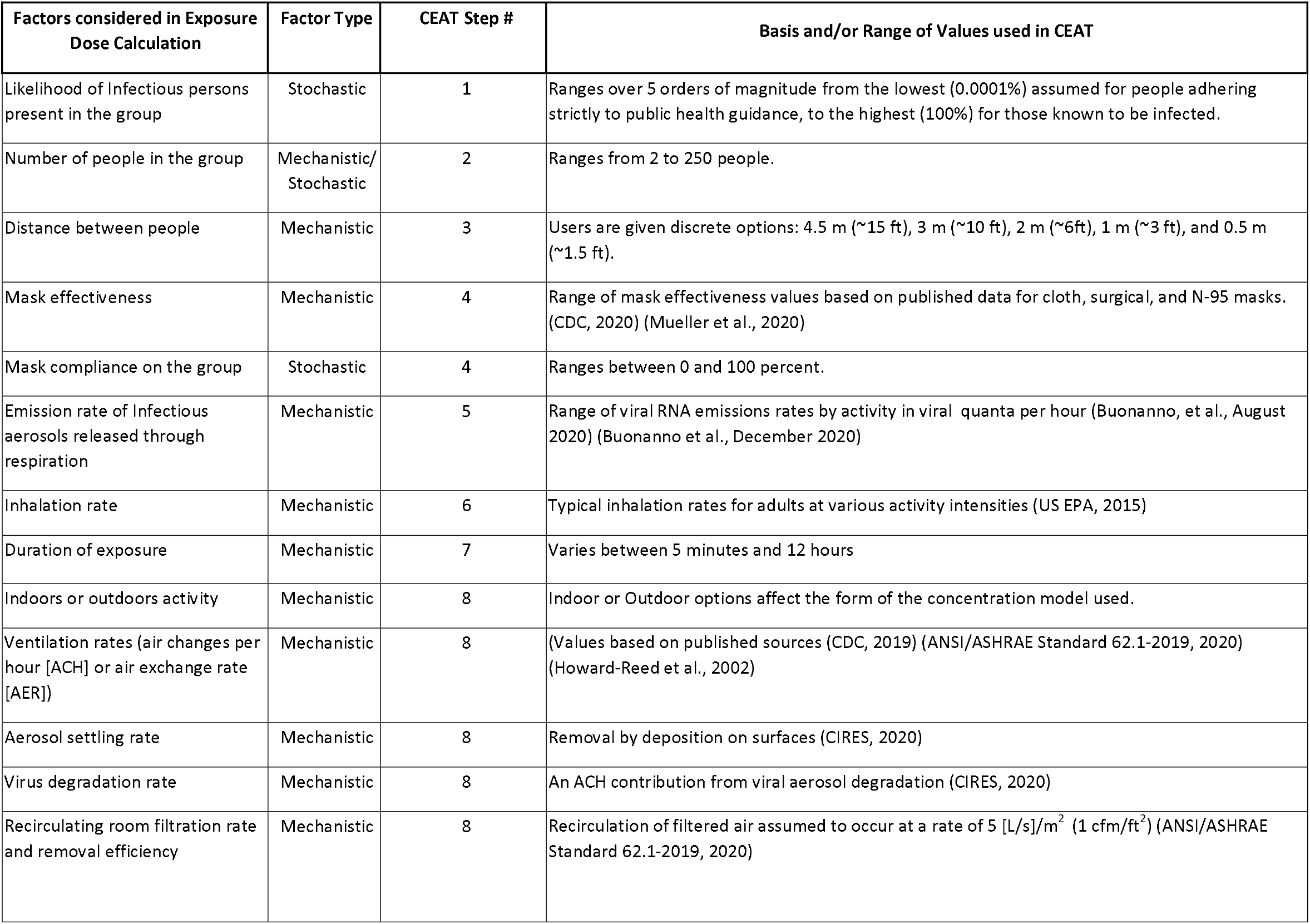
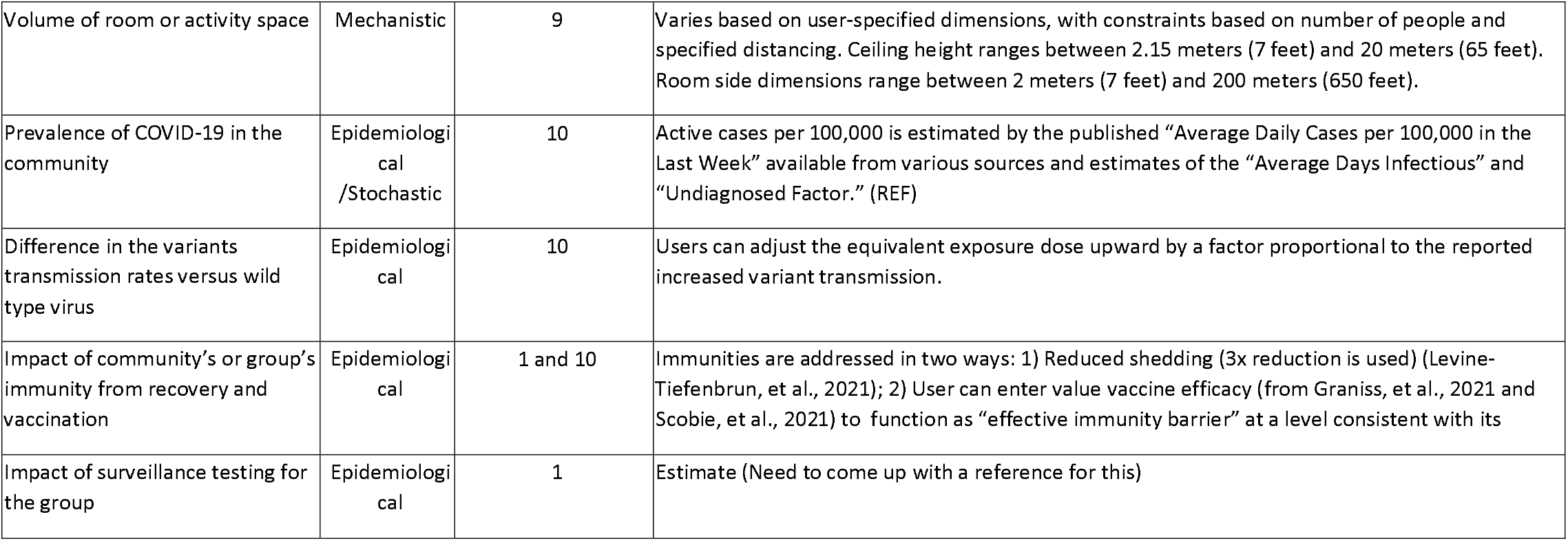
Summary of Factors. Mechanistic and epidemiological factors included in the Nomogram Model that affect exposure and inhalation dose.

Exposure is defined as the contact of an agent with an external boundary of a receptor (exposure surface) for a specific duration [29]. Dose is the amount of material that passes through the boundary based upon the intake rate, concentration, and exposure time. In this case, the boundary is the entrance to the respiratory system (i.e., through the nose, mouth, and other mucosa) [29] and the intake rate is the inhalation rate. Importantly, rather than a mass of material, we are only concerned with the quantity of material that contributes to transmission of disease. For viral dose-response models, the disease-causing quantity is often expressed in plaque forming units (PFU). A Wells-Riley based model expresses dose as an amount of quanta and when applied in a Poisson probability distribution, the complement of the Poisson distribution’s probability mass function (with the assumption number of occurrences is zero) can be used to predict an infection rate [15,30,31]. Engaging in activities with high inhalation and exhalation rates, such as group exercise, strenuous work tasks, or singing [32], is thought to correlate with higher doses and transmission risks [33]. Dose is the appropriate endpoint for a risk model, since it captures the contributions of concentration, exposure time, and inhalation rate. Since the model is meant to evaluate risks for events that include groups of people, and the number of people in each group is a variable that can be adjusted when planning events, we use a total group dose (*D*_*Group Dose*_) as the basis for our model:

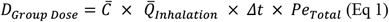

where 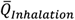 is the average inhalation rate for the group, 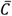 is the average concentration of the agent (in this case, aerosols containing SARS-CoV-2), ⊿t is the duration of group exposure, and *Pe*_*Total*_ is the number of people exposed in the group, which we will assume are all of the people in the group. The *D*_*Group Dose*_ represents the total quantity of infectious material that enters the respiratory tracts of all members of the group by inhalation over the duration of the potential event.

Rather than using an explicit calculation of group dose, the CEAT model takes the form of a relative dose model, comparing a specific evaluated scenario to a defined high risk baseline by a ratio:

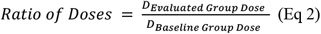

When the specific scenario results in a value that is equal to the baseline scenario, the ratio is The ratio may be orders of magnitude greater or less than 1 depending on the specific evaluated scenario. By benchmarking the dose calculations to a baseline scenario that is considered high risk by the Occupational Safety and Health Administration (OSHA), the model’s results can be aligned with the OSHA classifications of exposure risks [34] (see **Table S1**). The OSHA risk classifications depend on the industry type, the need for close contact (i.e., within 6 feet or approximately 2 meters) with people known to be or suspected of being infected with SARS-CoV-2, or requirement for repeated or extended contact with persons known to be or suspected of being infected with SARS-CoV-2 [34]. We define the baseline scenario to represent a person (perhaps a healthcare worker) who is exposed to a COVID-19 infected person for 15 minutes in an indoor setting with typical ventilation. We apply a range of assumptions to this scenario, addressing each of the factors in **Table S2** to arrive at a baseline scenario. This scenario was estimated to be consistent with between 4 and 9 percent likelihood of infection, based upon the range of infection rates reported in various studies due to close contacts, presumably involving wild type SARS-CoV-2 transmission based upon the dates of the cases included in the studies in early 2020 [35,36]. The inhalation dose values for other scenarios are compared to the baseline value through the simple ratio.

A critical variable that must be estimated by the model is the concentration of virus-containing aerosols that occurs as a result of the exhalation from people in the group at the event. The underlying concentration model used in CEAT assesses both the contributions of concentration due to the proximity of people (i.e., people in the “ near-field” whether indoors or outdoors) and the buildup of concentration in a room over time (i.e., “far-field”) after Nicas, 2009 [37]. As presented in the **STARS Methods**, to determine the near-field concentration, we employ a method that captures the effect of turbulent mixing that occurs due to higher air changes through ventilation or increased mixing of the air (e.g., through the HVAC system recirculating the air). Specifically, we employ equations that use the air change rate and total volume for an indoor space to calculate an eddy diffusivity based on relationships previously proposed [38–40]. We apply the calculated eddy diffusivity in a novel way that still allows us the advantage of using the computationally simple near- and far-field approach. Outdoors, only the near-field concentration contributions are used since the far-field is considered to be negligible [41]. Furthermore, also presented in detail in the **STARS Methods**, the CEAT concentration algorithm uses an approach for extending the near- and far-field approach to groups of people at set distancing intervals through the application of the superposition principle [42]. The superposition principle has been applied in the modeling of outdoor air pollutants from multiple sources [43] and to indoor air quality modeling of gaseous pollutants [38].

An important driver of the dose calculation is a stochastic approach that estimates the expected value of the number of infections in the group, since this is correlated to the quantity of virus-containing aerosols emitted in the modeled scenarios. In the baseline scenario, we assumed there to be one infected person. For the evaluated scenario, the number of infections in the group is dependent on the user’s estimate of the group’s behavior characteristics and an estimate of the number of active cases in the community population, calculated using the prevalence of diagnosed COVID-19 in the community, an estimate of the duration of infectiousness, and an estimate of the fraction of cases thought to be undiagnosed. The resulting number of active infections may be less than or greater than 1.

We adjust the calculated dose ratio result by additional factors: 1) concentration of virus-containing aerosols that occurs as a result of the exhalation from people in close proximity, 2) number of infections in the group, 3) current community prevalence of variants, 4) relative infectiousness of the prevalent variants; 5) current prevalence of immunity in the community of group gained by recovery or vaccination, 6) efficacy of immunity in preventing transmission, and 7) efficacy of surveillance testing of the group. The full CEAT dose ratio equation (**Eq. 3)** is shown in **Fig. 1B**, along with a mapping of where each of the CEAT step’s inputs are applied in the equation. The expanded version of the CEAT dose ratio equation, showing the NF and FF terms is found in Equation S46.

Users can use the tool to assess two side-by-side scenarios, and results are shown for the worst-case individual dose ratio, total group dose ratio, and near- and far-field contributions to the total group dose ratio. In the CEAT tool user interface, we refer to this dose as an “exposure” rather than a “dose,” since exposure is a more recognized term and will not be misconstrued by a user to have any association with a vaccine dose or medication dose. The group dose ratios for both scenarios are then categorized into four exposure risk bins, ranging from “Lower Exposure” through “Very High Exposure” and presented graphically (**Fig. 1A**). The model’s results include:

- **Group-wide Exposure (Dose) Ratio (Fig. 1B, Eq. 3):** Ratio of the group-wide dose to the baseline group-wide dose. This result is also shown in the bar graph. This result takes into account the dose that group members are exposed to, as well as the size of the group. Accordingly, this group-wise result provides an evaluation of the overall risk of the event.
- **Far-Field Group-wide Exposure (Dose) Ratio:** Portion of the group-wide dose that is due to the well-mixed concentration in the room.
- **Near-Field Group-wide Exposure (Dose) Ratio**: Portion of the group-wide dose that is due to the localized concentration in the room due to the proximity of people.
- **Individual Exposure (Dose) Ratio (Fig. 1B, Eq. 4):** This is the ratio of the individual-dose to the baseline individual dose.
- **Individual Dose (Fig. 1B, Eq. 5):** An estimate of the highest-exposed person’s dose in units of quanta.
- **Infection Rate (%):** To determine the infection rate, the Individual Dose is applied to a Poisson distribution to calculate the probability that the exposed group will become infected. The estimated rate of infection within the group can be inferred from this probability. The relationship between the dose and the infection rate can be adjusted through using a variable in the model called the “Poisson Distribution Adjustment Factor” in Step 10, which provides a linear adjustment factor to that relationship. The dose is multiplied by 1 over the adjustment factor.
- **# of Index Infectors:** Provides the assumed number of the infected individuals that were present based upon the selections and inputs in Step 1, Step 2, and Step 10 in the model. The model used this value to estimate the initial source(s) of infection in the room or at the event. This value can be a less than one person or fraction of a person, since it represents a probabilistically-determined number of people

### Demonstration of the Model Applied to Documented Transmission Events

We have demonstrated the effectiveness of CEAT results to predict infection rates for known transmission events by assembling data from eleven transmission events that were documented in the literature (listed in **Table 2**). All of the events, except for one, occurred before the vaccines were available and before the emergence of SAR-CoV-2 variants. To 1evaluate each of these scenarios in CEAT, we collected the data needed for each step and set the average daily cases per 100,000, such that the “Number of people initially infected” in the results would be equal to one, assuming there was one index case in each scenario. There are two ways of conceptualizing how the CEAT model is addressing this scenario of a known infected person (or index case), which are both mathematically equivalent:

**Table 2.** Reported COVID-19 transmission events.

| Case Number | Event Description | Volume of Room or Facility (m<sup>3</sup>) | People at Event | Total Cases Attributed to the event (Secondary Cases) | Total Infected Total Susceptible | Primary Reference |
| --- | --- | --- | --- | --- | --- | --- |
| Case 1 | Bus, Zhejiang Province, China, 19 Jan 2020 | 80.0 | 68 | 23 | 34% | Shen, et al., 2020 |
| Case 2 | Restaurant, Guangzhou, China, 24 Jan 2020 | 480.4 | 89 | 9 | 10% | Li, et al., 2021 |
| Case 3 | Meeting, Munich, Germany, 21 February 2020 | 210.0 | 13 | 12 | 100% | Hijnen, et al., 2020 |
| Case 4 | Commercial Aircraft, Flight VN54 (London, UK - Hanoi, Vietnam), 1 March 2020 | 662.2 | 217 | 16 | 7% | Khanh, et al. 2020 |
| Case 5 | Recreational Squash Game, Maribor, Slovenia, 4 March 2020 | 458.5 | 2 | 1 | 100% | Brlek, et al., 2020 |
| Case 6 | Call Center, South Korea, 8 March 2020 | 3267.0 | 216 | 94 | 44% | Park, et al., 2020 |
| Case 7 | Choir Rehearsal, Skagit Valley, WA, USA, 10 March 2020 | 808.0 | 61 | 32 | 53% | Miller, et al., 2021 |
| Case 8 | Recreational Hockey, Tampa Bay, Florida USA 16 June 2020 | 14452.7 | 24 | 15 | 65% | Atrubin, et al. 2020 |
| Case 9 | Restaurant, Jeonju, South Korea, 17 June 2020 | 184.8 | 13 | 3 | 25% | Kwon, et al, 2020 |
| Case 10 | Court Room, Vaud, Switzerland, | 149.5 | 10 | 4 | 44% | Vernez, et al., 2021 |
|  | 30 Sep 2020 |  |  |  |  |  |
| Case 11<br>(Omicron) | Holiday Party,<br>Oslo, Norway, 30<br>Nov 2021 | 1062.7 | 111 | 80 | 72% | Norwegian<br>Institute of<br>Public<br>Health, 2021 |

1. A receptor at the center and all others are potential sources. The emissions may occur from any one of the sources. We calculate an expected value of the dose for the person at the center, assuming that all of the people are equally likely to be the emitter, with a probability of *φ*, where *φ* = 1/(Number of People-1)) and the one emitter has a quanta based emission rate of 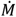. CEAT sums the results from all people (both FF and NF), s and multiplies by *φ*. This is the expected value of dose that the person located at the center of the group would receive if there was one emitter in the room, given that the emitter could have been anywhere in the room.
2. The source is at the center and all of the people are receptors. We calculate the expected value of the dose for each receptor (i.e., each susceptible person) given an emitter at the center, emitting at 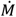. CEAT sums the results from all people (both FF and NF) and then divides by the number of receptors (i.e., number of people - 1) to arrive at an average. The result is the expected value of the mean dose that all people would receive in the room from the one emitter.

When examining CEAT performance for transmission events, we use the event’s number of infections, and the infection rate, *P*, which is the number of secondary cases (total infected) divided by the total susceptible people, yielding, 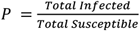. The total number of total susceptible is equal to the total people minus the index cases (typically equal to one). Using the information describing the event’s duration, room size, ventilation rate, activity type and any information on the location or the spacing of the people (**Table 2**), the CEAT individual dose is calculated. The infection rate can be predicted using the same statistical approaches that are used in the Wells-Riley model [30], in which the probability of at least one infection is computed using an assumed Poisson distribution, with shape parameter equal to *D*_*CEAT i*_, as shown in **Eq. 7**:

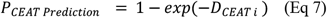

The CEAT predicted infection rate is plotted against the observed cases among the susceptible people (**Fig. 2A**). Information on vaccination, variant, and mask usage (which was none) was gathered from the reported events (**Table 2**). The CEAT results show a high correlation with the observed with an almost 1-to-1 (i.e,. R^2^ = 0.94) relationship to the observed infection rates (**Fig. 2A**). In addition, CEAT correctly binned the events as high risk and there is a significant positive correlation between the number of observed infections and CEAT group-wise dose ratio (**Fig. 2E**). As discussed in the previous section, to assess infection rate the initial relationship between the dose and the infection rate is unadjusted and then through the “Poisson Distribution Adjustment Factor” in Step 10 we achieve the corrected adjustment. With CEAT, even before this adjustment takes place, we still observe a strong correlation to the observed infection rates (**Fig. 2B**).

As a comparison to the CEAT results, the traditional Wells-Riley result that assumes a well-mixed dose, 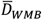, is calculated using:

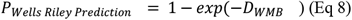

Using the same assumptions applied to the CEAT model for each of the events, including the same quanta-based emission rate, the Wells-Riley predicted infection rates for both the adjusted (**Fig. 2C)** and unadjusted (**Fig. 2D**), clearly show poor predictions when compared to the observed infection rates. The CEAT approach clearly outperforms the Wells-Riley in predicting infection rate in these cases (**Fig. 2E**). Interestingly, we also observe that the CEAT outperforms the Wells-Riley model with several other important parameters which include: distancing, density, breathing rate, and volume of the room (**Fig. 2E**).

### CEAT use to determine risk assessment for social gatherings

To demonstrate how CEAT can estimate potential exposure risk to COVID-19 for gatherings and events, we used CEAT to assess a set of hypothetical gathering scenarios that could have occurred in three locations in the United States (**Fig. 3**) using published CDC county-level COVID-19 7-day average new case data for the locations on 31 January 2022 [44]. We chose three representative locations: 1) a county with a low vaccination rate and high 7-day average new case rate (Knox County, TN), 2) a county with a moderate vaccination rate and a 7-day average case equivalent to the national average (Suffolk County, MA), 3) a county with the high vaccination rate and low daily cases (Montgomery Country, MD). At the time of analysis for all counties the Omicron variant accounted for >99% of COVID-19 cases [45]. We assumed the gatherings lasted 5 hours and would be held both indoors and outdoors. We also included a range of scenarios for distancing, type of masks being used, composition for the group of people, and location (i.e., indoors or outdoors). Lastly, we included analysis for 3 different group characteristic scenarios for the gatherings: 1) the general public (i.e., “equal to the community average”); 2) groups of people that are 100% vaccinated and follow all public guidelines; and 3) groups of people that are 100% vaccinated, follow all public guidelines, and testing was required before the gathering.

**Figure 3.**
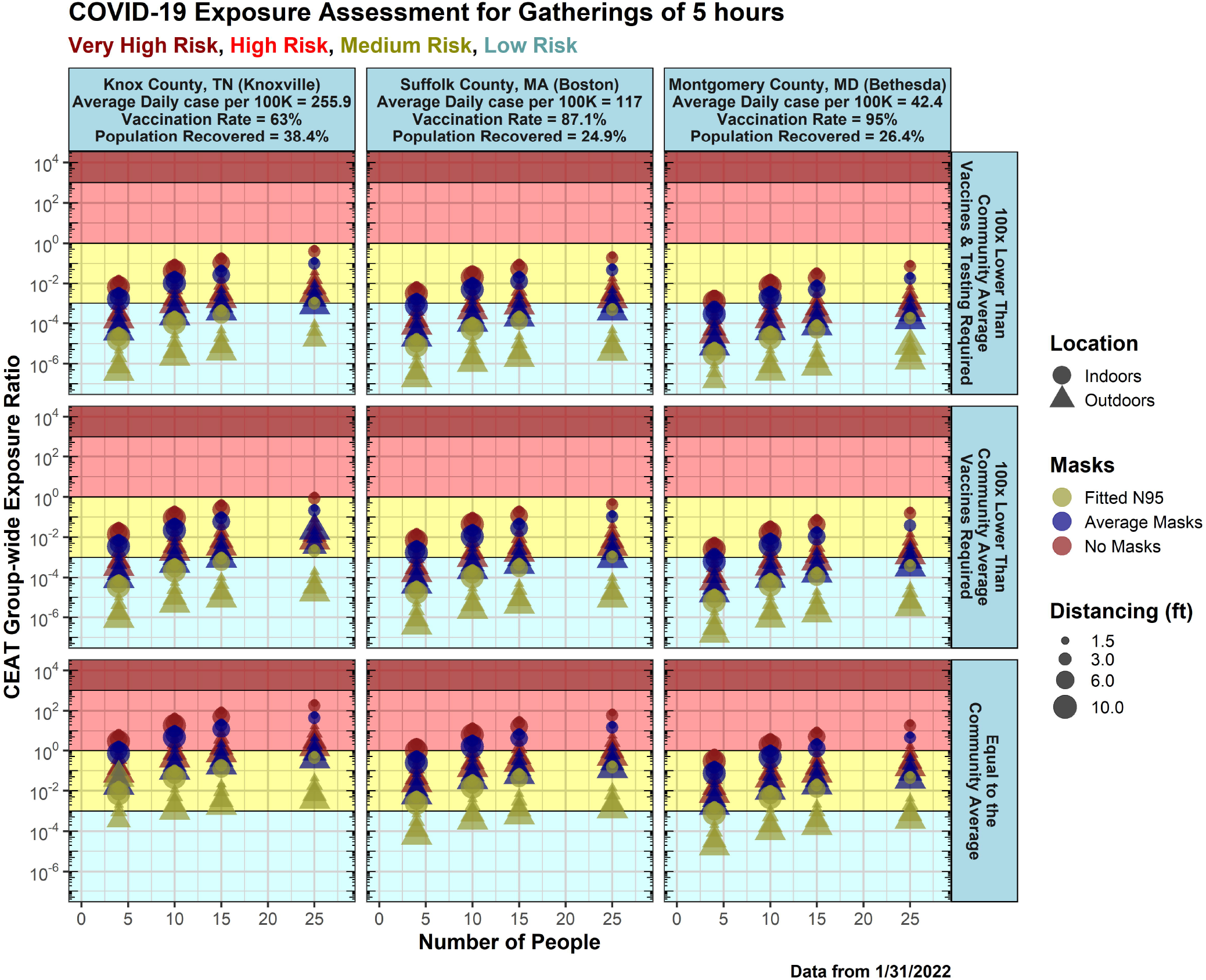
COVID-19 Exposure Assessment for Gathering Lasting 5 Hours. Data was analyzed on January 31st, 2022 for three US counties from the lowest (Montgomery County, MD) to highest (Knox County, TN) COVID-19 cases. The time was kept constant for all data points which assumes an average gathering of around 5 hours. The vaccination rates and population recovered rates are displayed on top of the plot for each county. Different scenarios were represented for location (outdoors = triangle, indoors = circle), distancing (increasing point size relates with increasing distance), and mask usage (no masks = red, average masks = blue, and N95/KN95 = yellow). The background shading of the plot indicates whether the data points are considered low risk (light blue), medium risk (yellow), or high risk (red) for COVID-19 exposure.

The exposure assessment from this analysis can help guide individuals to safely plan gatherings and events. As expected in all scenarios, if the gathering is composed of 100% vaccinated individuals that were tested and follow all public guidelines, the exposure risk is very low both indoors and outdoors, with the best masks and with an increasing number of people it increases to medium risk (**Fig. 3**). When considering the gathering in a general public scenario (e.g., eating at a restaurant), for all indoor scenarios in all counties without a mask with >10 people in the room, the group is at high risk of exposure to SARS-CoV-2 even when spaced 3 meters (approximately 10 feet) apart. Overall, we demonstrate with CEAT the more precautions are followed, the greater the reduction of exposure. This approach could be used as a guide for the public on how to use CEAT to properly determine the safest way to assemble while keeping the risk of exposure to COVID-19 low.

### NASA Ames Research Center used CEAT to determine the safest method for allowing workers to return to work

CEAT has been used by the NASA Ames Research Center (ARC) safety office to assist them in planning for workers to return to their campus. Starting on December 11, 2020, NASA ARC began to use the first beta version of CEAT to assess whether the tool could assist in gaining understanding on how to keep essential workers safe when having to work in person on the NASA ARC campus. To demonstrate how NASA ARC safety office has utilized this tool, we provide their assessment of exposure potential in 73 different scenarios throughout the campus (**Fig. 4** and **Data S1**). Since NASA ARC has been using this tool throughout the pandemic, every assessment used the latest COVID-19 case numbers from the State of California [46]. As is shown in **Fig 4**, the case numbers will vary due to the changing number of cases for that particular date of assessment, so it is essential to analyze the risk continuously with the most up-to-date COVID-19 case rates.

**Figure 4.**
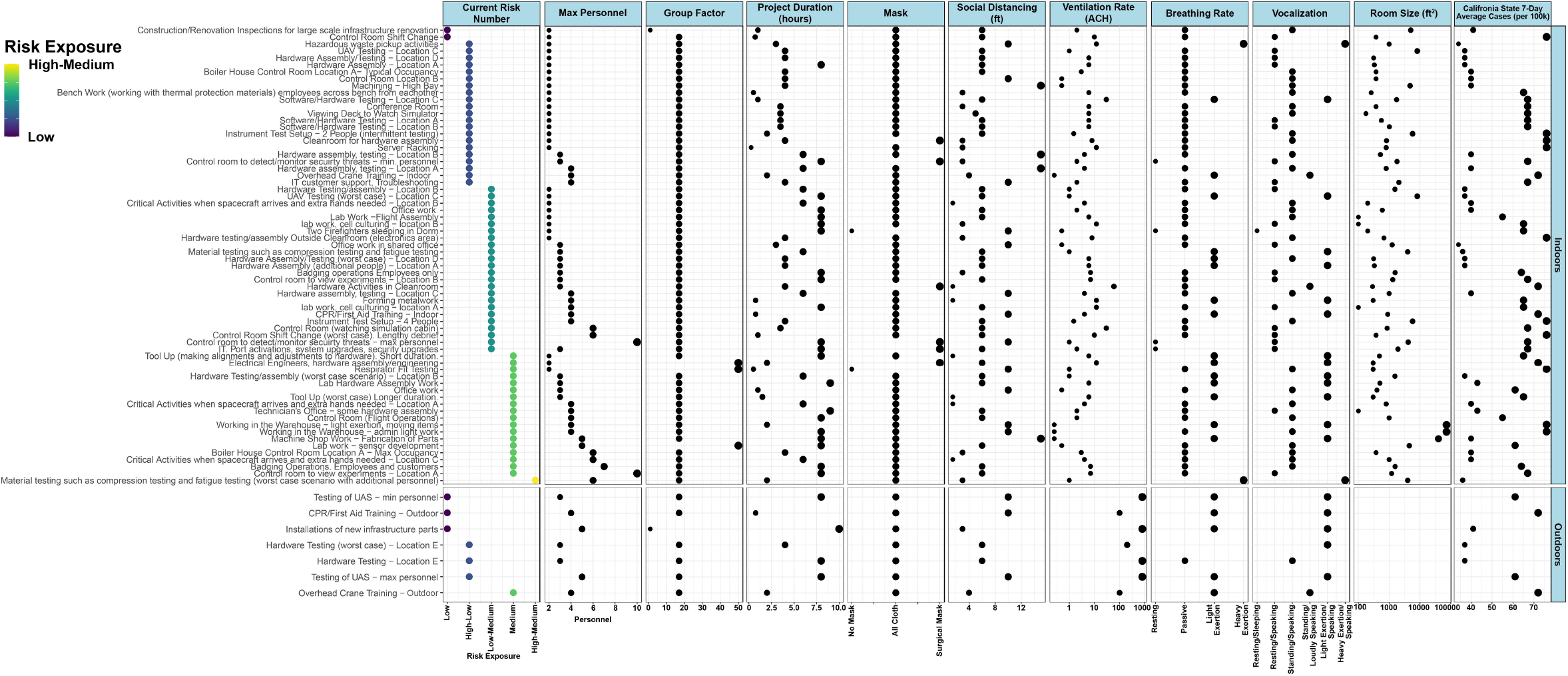
COVID-19 Exposure Assessment for Determining Lowest Exposure Risk for In-Person Work by NASA Ames Research Center. Exposure risk ratios using CEAT were calculated for 73 different scenarios (i.e., various locations and operations) at NASA ARC. The variables used for all ten steps are depicted for each scenario highlighting how various inputs affect the exposure risk ratios. The data for this figure is available in **Data S1**.

For each scenario, CEAT was used to determine the maximum number of personnel that could be allowed to be in each location such that the exposure risk was the lowest, while still allowing the work to be performed (**Fig. 4** and **Data S1**), which during pre-pandemic would have been occupied by more personnel. These maximum occupancy numbers were included in the project’s Return To Onsite Work (RTOW) plan that was reviewed by the safety office. In general, most operations could occur with one to two people thus reducing the potential exposure and resulting in a lower exposure risk. However, some operations required up to 10 personnel to be fully functional. As expected, these conditions increased the COVID-19 exposure risk to medium level. NASA ARC considered the group of people at work to be composed of people following all public health guidance which had the effect of reducing the assumed probability of COVID-19 prevalence in the group below the average for the community, with exception of a few locations where employees from organizations outside NASA ARC would participate. Social distancing was assumed to be the maximum possible for that work to be performed. For some locations such as “Critical activities when spacecraft arrives and extra hands needed - Location C” social distancing could not be achieved while performing the work, so other factors were considered, such as limiting the project duration, to find the lowest risk exposure estimate possible for that location and operation.

The breathing rate and vocalization for each location were also part of the decision in determining the maximum number of people in each location. Of all the locations and operations analyzed, only one location/operation resulted in the worst-case scenario which produced the highest risk exposure assessment (i.e., “High-Medium Risk Exposure”). The operation “Material testing such as compression testing and fatigue testing” typically involved high exertion physical activities as well as heavy exertion for the breathing rate and speaking over a long duration. The majority of the other locations and operations only required passive breathing rate and standing/speaking. The various operations that required elevated breathing rate and vocalization to light exertion typically had shorter project durations in order to reduce the COVID-19 exposure risk.

To provide inputs for the Air Changes per Hour (ACH), the ventilation rates for each location were either provided by the building managers, were directly measured, or were assumed using the guidelines in Step 8 in the CEAT. The most accurate ventilation rates available to the safety office were used in the model for each scenario. With the available data and estimated parameters in some cases, CEAT allowed NASA ARC to determine the operation-specific mitigation approaches, allowing its essential workers to return to work in-person with the low exposure risk to COVID-19.

CEAT has also been effective in allocating project resources and PPE where they would be most beneficial. When reviewing RTOW plans, NASA ARC safety office used the CEAT as a resource to recommend whether limited KN95/N95 masks would be effective at reducing potential exposure risk. Similarly, projects used CEAT when purchasing portable air cleaners (PACs), calculating the number of ACH needed to reduce risk to acceptable levels, typically “Lower Exposure”. Multiple projects found the number of PACs needed to reduce risk to acceptable levels were not financially feasible, and other controls such as increasing mask effectiveness and/or working in a different location were more cost effective for the same risk reduction. This allowed projects to spend their budgets more efficiently.

When the workplace face mask policy became optional for vaccinated personnel, CEAT was used to identify potential locations and operations where face masks would be required regardless of vaccination status. Personnel working in locations and/or operations where the relative exposure risk was in the “Medium” or “High” category were required to wear face coverings regardless of vaccination status. CEAT was especially effective in this regard as it allowed the safety office to provide this guidance using a consistent and unbiased method.

When tracking the CEAT model results over time, one can examine how the model responds to changes in community conditions and changes in organizational policies. NASA ARC tracked their worksite-specific relative group-wide exposure ratios with the California seven-day case rate (**Fig. 5**). There was a strong correlation (correlation coefficient=0.9759) between the two results, as would be expected, since the seven-day case rate is an input into the CEAT model in Step 10 (**Fig. 1B, Eq. 3)**. Specifically, NASA ARC used the location- and operation-specific exposure risk ratios that were assessed on a biweekly basis to calculate a “Centerwide Accepted Median Exposure Risk Ratio”. The fact that the CEAT results moved up and down with the community conditions allowed the NASA ARC safety office to adjust its guidance and mitigation strategies accordingly.

**Figure 5.**
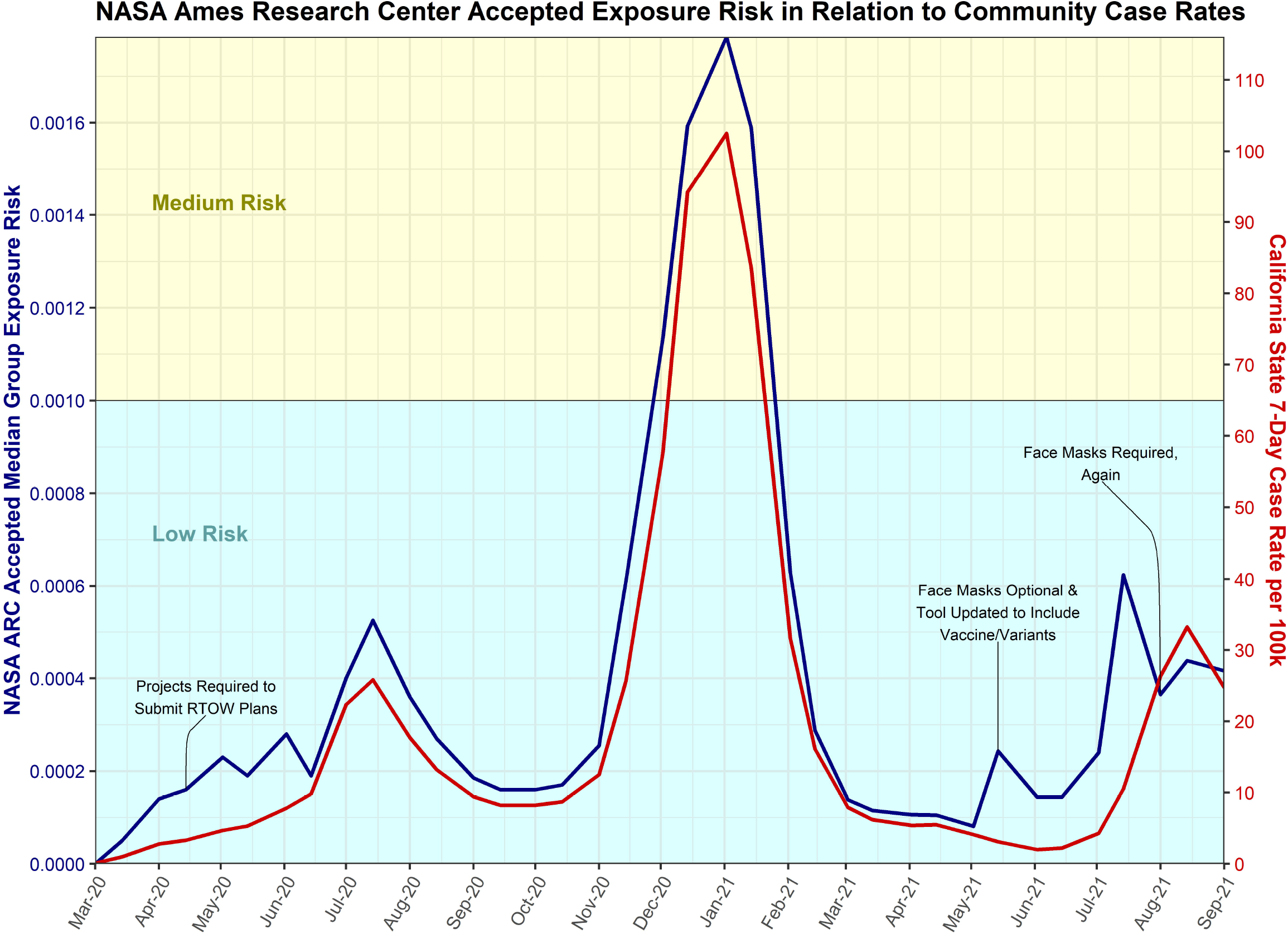
NASA Ames Research Center (ARC) Accepted Exposure Risk in Relation to Community Case Rates. Exposure risk ratios were calculated on a biweekly basis for 76 different scenarios (i.e., various locations and operations) at NASA ARC starting March 1, 2020 upon approval to RTOW through September 1, 2021. Biweekly reassessments included changes in community conditions such as case rate, variant prevalence, and vaccination rates in California. The median of all projected exposure risk ratios was calculated on a biweekly basis to establish a “NASA ARC Accepted Median Exposure Risk” (blue). These values were plotted along with the California state 7-day case rate per 100 thousand (orange). Notations were made designating major events and/or policy changes that may have influenced trends and deviations. The background shading of the plot indicates whether the data points are considered low risk (light blue) or medium risk (yellow) for COVID-19 exposure.

Beginning May 14, 2021, NASA ARC implemented the updated CEAT that included variant prevalence. It was noted that the correlation between the “Centerwide Accepted Median Exposure Risk Ratio” and the California case rate immediately decreased. However, at the same time this updated CEAT iteration was implemented, the NASA ARC face mask policy became optional for vaccinated personnel. Once NASA ARC reinstated their face mask policy for all individuals regardless of vaccination status, the correlation between “Centerwide Accepted Median Exposure Risk Ratio” and the California case rate appeared to return to similar values before May 14, 2021.

## Discussion

By establishing a set of equations that included mechanistic factors affecting near-field and far-field concentration, filtration, group behavior, and SARS-CoV-2 infection and immunity prevalence in the community, we developed a flexible, simple-to-use COVID-19 Exposure Assessment Tool (CEAT). The tool achieved our goal of allowing businesses, schools, government agencies, and individuals to assess COVID-19 exposure to the risk for groups and organizations. The tool is easy to use, computationally fast, and built on a well-developed and documented mathematical model that includes aerosol behavior, knowledge of SARS-CoV-2 transmission dynamics, as well as the effect of proximity. Here, through our comparison of CEAT results against observed transmissions for documented events, we demonstrate that CEAT can provide the accurate predictions compared to known cases with observed infection rates (**Fig. 2**), an example of how CEAT can be used for gatherings (**Fig. 3**), and real-time usage of this tool demonstrating how the NASA ARC safety office has, over the past year since the tool’s original release in December 2020, been evaluating how to allow essential employees to work in person with the lowest possible risk to COVID-19 exposure (**Fig. 4**).

As the CDC notes, the inhalation of fine respiratory droplets and aerosol particles is the primary means of SARS-CoV-2 transmission [47]. This is in line with recent publications that have shown that SARS-CoV-2 is spread by airborne transmission through the aerosols produced from breathing, talking, and singing [16,48–50]. Given this, the CEAT’s mathematical model addresses the aerosol dynamics and transport, treating the suspended aerosols as if they are dispersed as gasses would be using a eddy diffusivity approach, but also addressing deposition as a sink, using the same approach for aerosol deposition that was presented in the COVID-19 Aerosol Transmission Estimator spreadsheet-based tool [17], where an aerosol deposition factor of 0.24 hr^−1^ was used.

To demonstrate the model, we examined eleven literature-documented events with one of the events occurring in 11/2021 with known the Omicron variant and vaccine data [51] (**Fig. 2** and **Table 2**). When comparing CEAT results to the Wells-Riley model, CEAT better predicted the infection rate compared to the observed infection rates reported (**Fig. 2**). In addition, the exposure scores for all events predicted a high risk of exposures, which correlates to what was reported for each of these cases.

Schools and universities that have opened to in-person classes have been able to maintain low to no COVID-19 cases by applying many of the mitigation methods that are included in the CEAT, albeit independent of CEAT. The scenarios included in our assessment of gatherings (**Fig. 3**) can be applied to such environments and seems to match the observations that are being reported by schools and universities. Known outbreaks or superspreader events related to school openings have been chiefly reported occurring outside the classroom environment, including events during spring breaks [52] or athletic-related events [53], where enforcement of specific guidelines to reduce spread were not implemented. Other COVID-19 models for school re-openings have also shown similar recommendations as CEAT [54–56]. Brooks-Pollock et al. [54] provided a stochastic transmission model based on social contact data and patterns of student mixing to determine the impact and risk of COVID-19 transmission for universities in the United Kingdom. Since their model only targets social patterns and behavior of students contributing to COVID-19 infections, the focus for their mitigation strategies was with reducing the number of students in in-person classroom settings (i.e., increase social distancing), reducing living circles for students, and included regular testing. Although their model only takes into account one parameter from our model, the social distancing measures are in agreement with the exposure risk assessment results from our CEAT analysis. There also have been two independent agent-based models developed to assess SARS-CoV-2 transmission [56] and COVID-19 cases [55]. Similar to the model described above, Phillips et al. [56] agent-based model focuses on SARS-CoV-2 transmission based on children’s household sizes in the Ontario childcare centers and school buildings. They have also included parameters to take into account classroom sizes, sibling influence, symptomatic and asymptomatic rates, and physical distancing. Hernández-Hernández et al. [55] presented an agent-based model that utilizes the students’ community network to predict spread of COVID-19 within a school setting. They also considered the following parameters: 1) status of COVID-19; 2) physical distancing; 3) viral load; 4) hygiene standards; 5) confined spaces; and 6) social interactions. As expected, both models demonstrated the importance of social distancing and following the proper guidelines to prevent spread of COVID-19 which is in agreement with our model.

The recently published model by Miller et al. [57] utilizes CDC’s COVIDTracer Advanced tool to provide a transmission model for SARS-CoV-2 in schools. They took into account scenarios for infection in the community and public compliance to CDC guidelines to mitigate COVID-19 spread. Similar to other models, they found that social distancing is key to reduce spread and that the COVD-19 community case rate is crucial when assessing exposure risk. Other models of COVID-19 spread in small colleges only consider similar parameters as the models described above [58]. Although all these models provide a good basis for predicting the optimal conditions for having in-person classes, they miss key parameters incorporated in CEAT that are important to determine the most effective and accurate assessment of exposure risk to COVID-19. The lessons learned from classroom scenarios can also be applied to other gatherings, such as family gatherings. We demonstrated that depending on the location and people’s behavior, there are scenarios which have low risk for viral exposure (**Fig. 3**). Currently, there are no models specifically focused on family gatherings, but the literature available confirms the risk assessment analysis that CEAT generates. Whaley et al. [59] reported an assessment of COVID-19 risk associated with social gatherings, specifically during birthdays. They utilized data from 2.9 million households from a large insurance database that included COVID-19 prevalence from January 1 to November 8, 2020, and household birthdays across geographical regions in the US. They estimated that increased cases of COVID-19 correlated with social gatherings (i.e., birthdays), with increased cases for households in counties with higher COVID-19 prevalence. This study can serve as a population confirmation for the assessment we provided for gatherings with CEAT (**Fig. 3**).

Since the beta version release of CEAT in December 2020, we have made several additions to the tool to improve it including: 1) an eddy diffusivity-based near-field concentration algorithm; 2) added an infection rate calculation, 3) accounted for new SARS-CoV-2 variants, 3) address COVID-19 surveillance testing for groups, 4) addressed immunity within the population gained from recovery or vaccination. These additional functionalities were added to adapt to new information that was available since the initial release and reflect the team’s increased understanding of the risk dynamics related to SARS-CoV-2 transmission.

To determine the capability of CEAT being used by an organization to safely regulate employees working in-person, we have provided an example of it being applied by the NASA ARC safety office (**Fig. 4** and **Table S3)**. NASA ARC adjusted parameters related to group size, duration of time, and ventilation for the project to bring the exposure level to the lowest possible risk. The analysis shows how NASA ARC continued to monitor the changing case numbers within the community and utilized CEAT to provide the safest possible scenario for essential employees to work in-person. The use of CEATby NASA ARC represents a blueprint for other organizations, businesses, and schools to use the tool to manage their organizations exposure and risks to allow the organization to optimize mitigation strategies for employees to work in-person with lower exposure risk to COVID-19. The case study at NASA ARC has shown that as the push for employer vaccine mandates increases [60], employers can calculate the potential exposure risk reduction among their workforce compared to the community. This will be especially useful for workplaces in communities with low vaccination rates, where an employer vaccine mandate could have a large reduction in risk.

There are certain parameters in CEAT’s model that will have a greater influence on the assessment for exposure risk to COVID-19. As the examples we have provided show, the location of the gathering makes a big impact on the outcome (i.e., indoors vs outdoors) (**Figs. 3** and **4**). This is due to the fact that the exchange of aerosols between people outdoors will obviously be greatly reduced due to open circulation of the air versus in a confined space. The compliance to public health guidance policies is another parameter that will greatly change the outcome of the exposure risk assessment (**Figs. 3** and **4**). Lastly, the type of mask will make a considerable difference on the outcome of the model. From existing literature on effectiveness of masks [61–63], our model takes into account that the better the mask and the greater the adherence to mask wearing results in a reduction of dose ratio. (**Figs. 3** and **4**).

We believe that this tool and model can be easily modified and applied for guidance in current and future epidemics/pandemics from respiratory pathogens. In addition to SARS-CoV-2, a systematic review of the literature has shown that measles, TB, chickenpox, influenza, smallpox and SARS have strong and sufficient evidence of an association between their transmission and ventilation and air movement [64]. Accordingly, if pathogens have similar transmission mechanisms through aerosols, CEAT model can be modified to include the aerosol and viral dynamics to accommodate their pathogen-specific exposure risk assessment. We believe that by providing CEAT to the general public and building on its capabilities, will have a long-lasting beneficial impact for both our current pandemic and many other scenarios.

### Limitations of the Study

As with any mathematical model, there will be parameters that cannot be fully captured. Our model has a number of inherent limitations; nonetheless, it has been demonstrated to be used to accurately predict COVID-19 exposure risk for a limited number of eleven documented transmission event cases. In some of these cases key parameters were not available but were instead estimated. A full validation of the model is needed where a separate complete training set and test data set were compiled and applied to the model validation.

Several simplifying assumptions were made in the development of CEAT that resulted in more conservative results. One conservative assumption was using the highest exposed person to represent all people in a group. In future versions of CEAT, we may calculate location specific exposure values that account for the location of each person in the space, versus every other person in the space. This approach would result in a lower group exposure estimate than we currently calculate.

The CEAT concentration model assumes exhalations behave isotropically (i.e., they disperse equally in all directions), are non-buoyant, and are continuous exhalations. In reality, exhalations are more complex and may range from violent expiratory events or more regular puffs [18]. Exhalation plumes are typically anisotropic jets and have a buoyant nature due to their relative warmth and higher humidity. CFD modeling has captured these dynamics [26–28]. To compensate for the fact that CEAT assumes plumes are non-buoyant, which likely results in CEAT over-predicting concentrations at breathing heights, we adjust the height of the near-field volume to be equal to the distance between the source and the receptor, mixing the emission in all directions and in a larger near-field volume. The anisotropy is more difficult to capture in a model, given that people in groups may be facing in different directions at any point in time and in some events, such as a classroom, people may have a more uniform directionality. Additionally, the direction of the airflow in an indoor space is dependent on the flowrate characteristics of the ventilation system, geometry of the space, geometry and type of air vents, doors, windows, differential heating and cooling, other fans in the building, movement of people, and indoor/outdoor environment interactions. The simple approach used by CEAT to arrive at a concentration, certainly could be improved upon for any specific situations using CFD modeling, however the computational complexity and run times would greatly increase. Experiments that included high temporal and spatial measurement of CO_2_ from people in a variety of indoor conditions may be useful for testing and optimizing the concentration modeling approach used in CEAT. Experimentation, similar to the studies conducted by Vernez et al. 2021 [65], but using exhaled CO_2_ and inert aerosols as tracers, could be accomplished to compare with results obtained using CEAT’s algorithms to validate its concentration models and possibly calculate adjustments for the eddy diffusivity.

The aerosol deposition and virus decay behavior are simplistically handled in CEAT by adding additional terms to the air change rate resulting in a lower concentration since the effective ACH is increased. CEAT uses the same values for aerosol deposition and virus decay of 0.24 hr^−1^ and 0.63 hr^−1^, respectively, for all conditions, using the same values recommended in CIRES, 2020. The deposition and virus decay rates should vary based upon environmental conditions and the nature of the exhalation conditions (breathing, coughing, sneezing, singing, and speaking). In future versions of CEAT, exhalation-specific deposition rates and the environmental-specific decay rates (i.e., varying by humidity, temperature, and ultraviolet radiation) could be calculated [66–68].

The model could benefit from the incorporation of vaccine-specific values for the efficacy of a vaccine to prevent transmission, recognizing that this approach is an oversimplification of a very complex process. In our model, we currently have “Protective Effectiveness of Immunity” being considered as one universal number for the population being assessed. We believe that differences for efficacy between the vaccines can be averaged for the overall community. Our model also lacks the ability to incorporate the length of time that has passed since being vaccinated or previously infected. This might change the risk assessments since we now know that for both cases the levels of antibodies against SARS-CoV-2 reduce over time [69–71]. However, more research is needed to determine how this impacts the protection against SARS-CoV-2, which is the reason we have not incorporated this parameter into our model at the moment. Additionally, CEAT does not account for the buildup of viral particles between groups utilizing the same space one after another. This limitation will impact groups gathering in a room separately, but sequentially. One way to overcome this limitation is for groups to allow a certain amount of time between occupancy. Calculating the amount of time needed for a ventilation system to remove 99% of contaminants, like that provided by the CDC for infection control in health-care facilities [72], can allow groups to calculate their exposure risk ratio during separate but successive events. Similarly, groups can estimate their relative risk of back-to-back gatherings by adding the total duration of all meetings throughout the day.

## Data Availability

All data produced in the present study are available upon reasonable request to the author.

## Acknowledgments

The opinions expressed in this article are those of the authors and do not reflect the view of Signature Science LLC, National Institutes of Health, the Department of Health and Human Services, or the US government. The authors acknowledge and thank Signature Science’s Mr. Jim Gibson, of Charlottesville Virginia, for the development of certain graphics for the manuscript and for the website development for the publishing of CEAT. Additionally, the authors acknowledge and thank Mr. Ken Martinez of the Integrated Bioscience and Built Environment Consortium (IBEC) for his review of the draft manuscript and his enthusiasm for the CEAT.

## Funding Acknowledgments

B.S. and M.I. developed the CEAT concept, method, model, code and user interface, conducted literature reviews, and prepared this manuscript using solely Signature Science, LLC provided Internal Research and Development (IR&D) funding. S.D. was supported by the DOE Office of Science through the National Virtual Biotechnology Laboratory (NVBL), a consortium of DOE national laboratories focused on response to COVID-19, with funding provided by the Coronavirus CARES Act. A.B. is supported by supplemental funds for COVID-19 research from Translational Research Institute for Space Health through NASA Cooperative Agreement NNX16AO69A (T-0404) and further funding was provided by KBR, Inc.

## Author Contributions

Conceptualization: B.S.; Creation of Model and Tool: B.S. and M.I.; Methodology: B.S., M.I.; Formal Analysis: A.B., B.S., C.G., N.S.T.; Analysis with NASA ARC usage: B.F.H., K.D.C., N.B.N, A.R.D., A.B.; Investigation: A.B., B.S., M.I.; Resources: A.B., B.S.; Writing Original Draft: B.S., M.I. A.B., N.S.T.; Writing original draft for NASA ARC usage: B.F.H., A.R.D.; Review & Editing: A.B., B.S., M.I., C.G., S.D., A.R.D., N.S.T., B.F.H., T.A.; Figures and Visualization: B.S., A.B., N.S.T., for graphical abstract: A.B.; Funding Acquisition: A.B, B.S.; Supervision: A.B., B.S.

## Supplemental Figures and Tables

**Figure S1. The different box models for CEAT. A)** Factors Included in COVID-19 Exposure Assessment Tool Interface (CEAT). A summary of the factors and mechanisms affecting the comparative dose and exposure risks. **B)** Single Zone Well-Mixed Box Model. Basic box model assumes emissions are instantaneously well mixed. **C)** Near-Field (NF) and Far-Field (FF) Box Model. “Box-within-a-box” approach provides localized higher concentration in the vicinity of the source.

**Figure S2. CEAT Concentration Model Performance. A)** Modeled results vs. measured concentrations using CEAT concentration model in cases when ACH was less than or equal to than 0.75 hr^−1^, applying the relationship between ACH, room size and eddy diffusivity according to Venkatram and Weil, 2021. Results are normalized by dividing by the emission rate. **B)** Modeled results vs. measured concentrations using CEAT concentration model in cases when ACH was greater than 0.75 hr^−1^ and assuming 4 vents per 100 meters^2^ of room area, applying the relationship between ACH, room size and eddy diffusivity according to Foat et al., 2021. Results are normalized by dividing by the emission rate. **C)** Modeled results vs. measured concentrations using CEAT concentration model for all cases.

**Figure S3. Dimensions and sources for the box model. A)** Near-Field (NF) and Far-Field (FF) Box Dimensions with Two People. **B)** The application of the principle of superposition with Near-Field (NF) and Far-Field (FF). **C)** Near-Field (NF) Triangular Prisms for the 1^st^ Ring of the Group. NF triangular prisms for the 1^st^ ring of the group that have a height and base of *D*_*tot*_, in a triangular grid with each side length of *D*_*tot*_. The first ring’s triangular prisms have a base area of A1 = ½ *D*_*tot*_^2^. **D)** Near-Field (NF) Triangular Prisms for the 2^nd^ Ring of the Group. NF triangular prisms for the 2^nd^ ring of the group that is equally spaced *D*_*tot*_. The second ring’s triangular prisms have areas A2 = ½ 2*D*_*tot*_^2^ and 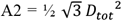. These average to *A*2 = 0.9330 *D*_*tot*_^2^. **E)** Source perspective with two sources shown. Under an assumption of isotropy assuming no predominate flow and a sufficient averaging period, the sources emit in all directions equally. **F)** The same two triangles that impact the receptor in the source perspective can be turned 180 degrees and are part of the potential set of triangles in each ring. The dimensions and parameters are identical between both views.

**Table S1. OSHA Risk Classifications**. OSHA’s classifications offer a means of comparing exposures to a scenario that can be defined as high risk [34].

**Table S2. Baseline Scenario Approach**. Mechanisms affecting the exposure risk.

**Table S3. Comparison of Three Simple Approaches to Modeling a Continuous Point Release**. In each of the three cases, the equations for concentration are nearly identical.

**Table S4. Triangular Prism Parameters and Equations for Each Ring**

**Data S1. NASA Ames Research Center (ARC) CEAT Data**. Calculated exposure risk ratios and CEAT assessment variables for different scenarios varied by location and operation at NASA ARC. The data in this figure is used in **Fig. 4**.

## Materials and Methods RESOURCE AVAILABILITY

### Lead Contacts

Further information and requests for resources and reagents should be directed to and will be fulfilled by the Lead Contacts, Afshin Beheshti and Brian Schimmoller.

### Materials Availability

This study did not generate new unique reagents.

### Data and Code Availability

The published article includes all datasets generated and analyzed during this study. Any additional information required to reanalyze the data reported in this paper is available from the lead contact upon request.

## METHOD DETAILS

### Relative Dose Ratio Approach and Exposure Risk Model Derivation

CEAT’s relative dose ratio approach is based upon a mechanistic dose-response framework. The starting point for the inhalation dose model is to use the relationship that defines group-wide inhalation dose as a linear system where:

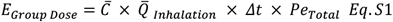

and 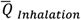 is the average inhalation rate for the 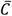 is the average concentration of the agent (in this case, aerosols containing SARS-CoV-2), ⊿t is the duration of group exposure, and *Pe*_*Total*_ is the number of people exposed in the group, which we assume is equal to the total number of people in the group. The *E*_*Group Dose*_ represents the total mass of contaminant that enters the respiratory tracts of all of the group by inhalation over the duration of the potential exposure or event. The fate or dynamics of the virus within the respiratory tract are not considered in the model and would be part of a transmission process. The critical variable that must be estimated by the model is the concentration of virus-containing aerosols that occurs as a result of the exhalation (i.e., breathing, speaking, coughing, singing) from people who are in close proximity and build up in a room over time.

There are a variety of ways of estimating concentration of contaminants in the air. Several commonly used methods include well-mixed box (WMB) models [73], computational fluid dynamic (CFD) models [74], and gaussian dispersion models [43]. Computational fluid dynamics based models uses numerical solutions of the first principle equations of fluid flow and contaminant transport that are tailored to the specific geometry, scale and temporal lengths, and flow regimes, and are capable of modeling the complexities of particle dynamics, inhalation, exhalation, and interaction with flows in a building [74]. Gaussian models use an explicit solution of the contaminant transport equations, and are, therefore, computationally fast compared to CFD models. Gaussian models are typically used at larger scale lengths (100s of meters or more) and are used in outside environments, not typically used in indoor modeling [75]. The WMB model is a simple model that can be used to estimate concentrations of contaminants in the air. It treats a room as if it were a continuous stirred-tank reactor (CSTR) and uses the basic equations for concentration that were developed for modeling continuous reactors in chemical engineering.

The WMB (or zone) approach is widely used, and, for example, is the basis for the National Institutes of Standard and Technology’s (NIST’s) CONTAM indoor air quality model [76]. NIST has also applied a single zone WMB approach in its Fate and Transport of Indoor Microbiological Aerosols (FaTIMA), where it assumes rooms are single well-mixed zones [77].

The basic equation for the single zone WMB is shown below:

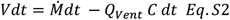

where V is the volume of the box, *Q*_*vent*_ is the ventilation rate (in units of volume per time) through the box, and 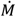 is the emission rate (in units of mass per time) (**Fig. S1B**).

If we assume the emission rate is constant starting at time equals zero, the time varying equation takes the form:

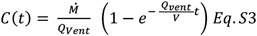

Once enough time has passed to achieve equilibrium, the model takes the simple form:

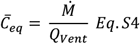

The basic simplifying assumption of the WMB model is that it assumes that a contaminant is instantaneously completely mixed throughout a volume of air. This instantaneously well-mixed assumption is a significant limitation when looking to determine the exposure between people in a room or space if they are in close proximity relative to the size of the room. The single zone well-mixed assumption results in the same exposure no matter how close or far people are located. Accordingly, methods that can assess the potential for higher concentrations (and exposures) that would result between closely clustered people would be useful for quantifying exposure, doses and associated risks.

### Near-Field (NF) and Far-Field (FF) Box Model

In the field of industrial hygiene, it is recognized that the single zone box model may underestimate exposures experienced by receptors (i.e., people) close to a hazard, since it assumes that the concentration is instantaneously well-mixed over the volume of the room (Jaycock et al., 2011). While computational fluid dynamics is one option to resolve the spatial complexity of dispersion and mixing of a contaminant, industrial hygienists have devised a simpler way of estimating the high concentrations near a source using a “box within a box,” with an inner box or near-field (NF) box containing the contaminant source and a receptor, and a larger, far-field (FF), box that represents entire volume (e.g., room). (**Fig. S1C**) The time dependent concentration at the receptor is estimated by adding the NF and FF concentration contributions [37,41].

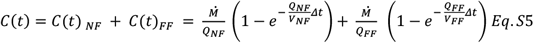

where, 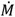 is the continuous mass release rate per minute of the contaminant of concern, *Q*_*NF*_ (or as referred to by Nicas as β) is the NF volumetric flow rate (m^3^ per minute), *Q*_*FF*_ is the FF volumetric flow rate (m^3^ per minute), *V*_*NF*_ is the NF volume (m^3^), *V*_*FF*_ is the FF volume (or volume of the room or activity space) (m^3^), and *Δt* (minutes) is the elapsed time since the start of the release.

If one assumes both boxes to be at equilibrium, the equation takes the simpler form:

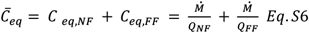

Calculating the ventilation rates, *Q*_*NF*_ (or as referred to by Nicas as β) and *Q*_*FF*_ using the room volume and appropriate air change rate specific for each volume yields:

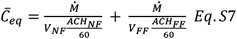

where *ACH*_*NF*_ is the NF air change rate (hr^−1^), and *ACH*_*FF*_ is the FF air change rate (hr^−1^). For the time dependent form, since the volumes in the exponential term cancel themselves out, the following results:

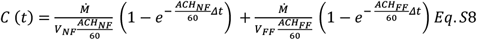

In the application of NF and FF models, it is recommended [37,78] that the NF flow rate, *Q*_*NF*_ (or β), be equal to 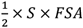 where FSA is the free surface area of the assumed NF control volume and S is a random air speed (instantaneous in random direction) at the interface of the NF and FF zones and 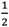 is used assuming that half of the air volume is entering the control volume and half of the air is leaving the control volume. Further, Nicas recommends using s=15.1 meters per minute (50 feet per minute) when strong air currents are present and s=3.0 meters per minute (10 feet per minute) when air currents are lacking near the NF zone [41]. A median random air speed for indoor office and home spaces was observed by Baldwin and Mayard, 1998 [79] to between 0.05 and 0.1 meters per second. Nicas, 2014 [41], referencing Baldwin and Mayard, 1998 [79], recommends that the typical value of 0.06 meters per second (3.6 meters per minute), may be used with the *FSA* approach in indoor settings. Accordingly, in *ACH*_*NF*_ can be calculated using the *FSA* approach as follows:

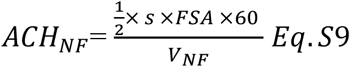

As we will show, the FSA approach when applied using typical values for median random airspeed did not predict concentrations that align well with measured data. Accordingly, we have devised an alternative way of calculating the *ACH*_*NF*_ using an effective value for the random air speed we call *s*_*eff*_ that varies with distance from the source and is derived from an estimate of the eddy diffusivity. To do this we examined the gaussian/ eddy diffusivity equations and show that the NF/FF equations can equivalent in certain cases to the continuous, gaussian solution of the dispersion equation when there is no advection (i.e., mean wind speed is equal to zero). Through this analysis, we can formulate a NF/FF model that uses eddy diffusivity rather than relying on the Baldwin and Mayard, 1998 reported random air speed to provide the mixing dynamics. To illustrate this, we start with side-by-side derivations for 1) a continuous point release using the gaussian approach, 2) the NN/FF model using a spherical NF volume, and 3) the NN/FF model using a hexagonal prism NF volume, as shown in **Table S3**. In all three cases, we arrive at equations for concentration that are nearly identical. Assuming the same values were used for *K* and distance from the source, all three representations would provide nearly the same result – even the hexagonal prism representation since 6 is within five percent of 2*π*.

The challenge in using an eddy diffusivity model is determining the appropriate value for *K* [80]. It is important to note that in the derivation of the gaussian solution, the value for *K* is assumed to be constant over the domain [43]. The form of the equation for *K* that we arrive at, *K* = *x* · *s*, is similar to the form suggested by Venkatram and Weil, 2021 [40], *K* = *α* · *u* · *l*. Venkatram and Weil, 2021 [40] describe *α* as a dimensionless value that would be determined experimentally; u is a representative velocity, and l was a representative length. For now we will assume that *α* = 1, such that in our case *K* = *D* · *s*.

Cheng et al. 2011 [38] show a relationship between the air change rate for a room and the eddy diffusivity using experimental measurements of carbon monoxide released in two indoor environments. The data from these experiments are presented in their paper and in Acevedo-Bolton et al. 2012 [81]. These approaches capture the additional turbulent kinetic energy that is added to the system through the higher air changes through ventilation or increased mixing of the air (e.g., through the HVAC system circulating the air) [38,40]. This approach provides for a constant eddy diffusivity within the room and does not suggest dependency of the eddy diffusivity on the distance from the source. The recommend the eddy diffusivity (m^2^ sec^−1^) is calculated using the mechanical *ACH*_*FF*_ (air change rate in hr^−1^) and, *V*, the overall volume of the room (m^3^), as follows:

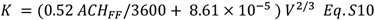

Venkatram and Weil, 2021 [40], using the same datasets suggest a more simple but similar relationship:

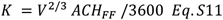

Foat, et al., 2020 [39] recommend a similar relationship that was arrived at through CFD simulations over a wide of range of indoor parameters:

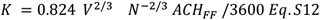

where *N* equals to the number of inlet vents for the room. Foat, et al., 2020 [39] looked at a range of room volumes between 50 m^3^ and 5000 m^3^, floor aspect ratios (length/width) between 1-3, height/(floor area)^2^ ratio between 0.1 and 1.5, and air change rate between 0.6 and 19.9 hr^−1^. In most modern buildings, the number of vents would increase with increasing volume or area. Across the range of 235 scenarios that were modeled, the number of vents per 100 m^2^ of area ranged from 3.8 to 8 vents per 100 m^2^ (excluding the four extreme values of approximately 50 vents per 100 m^2^), and averaged 4.6 vents per 100 m^2^. To limit the number of variables that the user needs to know or determine to use CEAT, we replace N with a relationship between the area of the room and a reasonable number of vents per unit area, examining values ranging from 3 to 8 vents per 100 m^2^.

By combining the two representations of the eddy diffusivity equations and assuming that the product of D and s is a constant, we can calculate an effective velocity *s*_*eff*_ (in m min^−1^) that is consistent with a constant eddy diffusivity at all distances from the source.

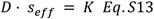

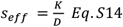

Below are the Venkatram and Weil, 2021 [40] and Foat, et al., 2020 [39] solutions for K expressed as *s*_*eff*_(in m min^−1^):

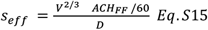

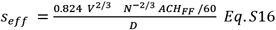

Since all three K and ACH relationships are constant with respect to D, the product of *D* · *s*_*eff*_ is a constant, thus any change in s is inversely proportional with the change in D. Therefore, as D increases moving away from a source, the value of *s*_*eff*_ decreases.

Now we come back to the relationship of *K* = *α* · *u* · *l* or expressed in our variables *K* = *α* · *s*_*eff*_ · *D* and define *α*, adjustments for the eddy diffusivity, as a means of capturing any dependency of K on distance from the source and adjustment to the dependence on ACH in the form (should measurement data indicate there are dependencies):

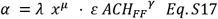

Substituting the equation for K into our original equation and rearranging so that the FSA of the hexagon is still calculated, we arrive at:

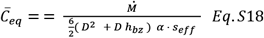

and *ACH*_*NF*_ is

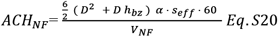

To calculate an average inhalation dose over a period of time, assuming that the initial concentration is zero (*C*(0) = 0) from a single source, we estimate the average dose by calculating the concentration at the midpoint of the duration, 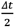.

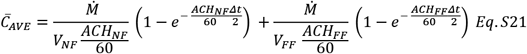

As the duration increases, the factors 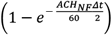 and 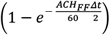 will converge on 1. Given that *ACH*_*NF*_ is likely greater than *ACH*_*FF*_ the factor 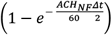 will converge faster than the factor 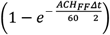. Meaning, the near-field term will achieve equilibrium faster than the far-field term.

### Validation the of the Single Source Equation with Measurement Data

We compare predictions calculated using Equation S20 to measurements of chemical and aerosol releases in indoor environments that characterize concentrations at various distances from sources. We included data from carbon monoxide (CO) releases in two homes [81,82], toluene releases in a test chamber [83], benzene releases in an industrial environment [84] and liquid aerosols containing lactose released to simulate an actual COVID-19 transmission incident that occured in a Swiss court room [65]. The study of carbon monoxide (CO) releases [38,81,82] occurred in two residential homes where seventeen separate 8-hour tests with continuous emission rates were conducted. Measurement distances from the source ranged from 0.25 meters through to 5 meters. The chamber tests conducted by Zhang, et al., 2007 involved simultaneous measurements at four points that were 0.1 meters from a release point. Across the dataset, the distance from the source varied between 0.1 meters and 5 meters, the room volumes varied between 3 m^3^ and 50,000 m^3^, and the air changes per hour between 0.17 and 218 hr^−1^. We also examined an example case that was presented by Nicas 2009 [37], where the conventional NF/FF approach is applied.

Examination of the performance of the three eddy diffusivity models (assuming that no adjustment is necessary and that value of *α* = 1 for the expression in **Eq. S17**) shows that the best model above 0.75 hr^−1^ is the Foat, et al., 2020 [39] equation with an R^2^ = 0.94 (**Fig. S2B**) when the number of vents per 100 m^2^ is equal to 4. The best model below 0.75 hr^−1^ is the Venkatram and Weil, 2021 [40] model, with an R^2^ = 0.92 (**Fig. S2A**). Acevedo-Bolton,et al. 2012 [81] show in their analysis that the carbon monoxide sensors (measuring at 15 second time intervals) at 0.25 meters were likely seeing concentrations that were above the upper limits of the instrument’s data logger (between 128 and 150 ppm), resulting in an underestimate of average reported concentration. Our model systematically overpredicts the concentrations, as compared to the measured data, at 0.25 meters and to a lesser degree at 0.5 meters. Consequently, we remove the 0.25 data from the dataset.

The major difference between the two models is the inclusion of a factor that captures the number of vents. It is reasonable to assume that spaces with very low air change rates do not have vents (or do not have functional vents), so the inclusion of the number of vents in the equation is not meaningful and in low air change rates the Venkatram and Weil, 2021 [40] equation is sufficient. Also, the lower limit of the air change rate in the Foat, et al., 2020 [39] dataset was 0.6 hr^−1^ and only three of the 235 modeled scenarios analyzed had air change rates less than 0.75 hr^−1^. Given that (1) most commercial and institutional facilities will have air change rates that are greater than 1 hr^−1^, (2) would have an additional air change rate term to account for HVAC recirculation and filtration, and (3) these facilities’ HVAC systems will include inlet vents, it is important to use a method that addresses the effect of vents on dispersion and is accurate at high air change rates. Also, given that the risks of COVID-19 exposure are highest when the air change rates are low such as when natural ventilation is relied upon, it is important to have a method that works well in those conditions. The Acevedo-Bolton, et al., 2012 [81] dataset and the Venkatram and Weil, 2021 [40] estimate for eddy diffusivity cover those low air change rate scenarios.

Based upon these factors and our analysis, in the CEAT model, we use the unadjusted Venkatram and Weil, 2021 [40] model to estimate eddy diffusivity at air change rates at or below 0.75 hr^−1^ and the Foat, et al., 2020 model [39] to estimate eddy diffusivity above 0.75 hr^−1^ with an assumption of 4 vents per 100 m^2^. The results of this combined model compared to the measured data, are shown in **Fig. S2C**.

### Multiple Sources

Equation S21 provides an estimate of the average concentration from one person’s emission at a receptor, but not the contribution of how multiple people’s emissions would affect the concentration. To address multiple sources in combination, the additivity property of the “superposition principle of linear systems” can be applied [42], which enables that the effect of each person’s emissions at a receptor can be calculated separately and summed. The superposition principle has been applied to outdoor air pollution dispersion modeling [43] and provides the theoretical basis for modeling complex scenarios involving multiple emission sources in outdoor gaussian plume models such as EPA’s AIRMOD [29]. The logic is, therefore, if a NF and FF approach can be used to estimate the higher concentration in the close proximity of one person to another person, then if *n* people were added to the system, and *n* additional NF boxes were added, the terms would be added to the equation for each person (i.e., emission source), with each being independent and summing to total concentration, as shown below:

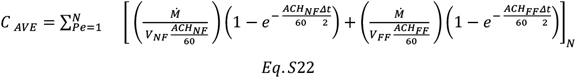

The superposition principle also includes a homogeneity property, which allows us to apply a scalar factor across all emission sources resulting in the concentration at the receptor changing proportionally to the value of the scalar. This property provides the conceptual basis that allows one to conclude that if the emission rate from each source is increased or decreased by a factor, that one could assume the concentration would increase or decrease by the same factor. The scalar could also be the product of several scalars, including a probability factor. CEAT will use this property defining a scalars, 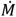 and *φ*(phi), to adjust both the emission rate and the probability of the emission rate, assuming that the emission rate and the probability of emission rate are constant for all sources for a given scenario, resulting in the following equation:

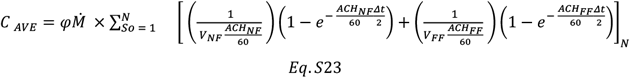

In a two-source system with one receptor, where the distance between the two receptors and the source equidistant shown in **Fig. S3A**, the following equation can be written:

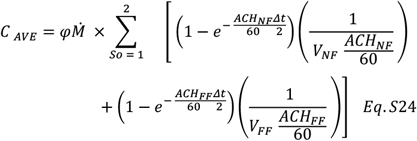

Using a hexagonal prism for the NF volume allows one to place the system of equations on a regular grid of equidistant triangles **(Fig S3A**). Using a regular grid of equidistant triangles, as compared to a regular rectangular grid, has advantages since all nodes are equidistant from their nearest neighbors. This equidistant neighbor feature is particularly useful given the objective to assess various distancing options. The use of a triangular grid allows one to conveniently draw a hexagonal prism that is made up of six triangular prisms that approximates a cylinder, with each centered on the six closest nodes to the receptor, **(Fig. S3B**). The orientation of the triangular prism within the box makes no difference to the calculations. Accordingly, we can rotate each of the triangular prisms 180 degrees for visual convenience (**Fig. S3C**). We do this because we can define the system identically from two perspectives, the source view and the receptor view.

The system shown in **Fig. S3D** can be used to evaluate both the NF and FF concentrations from up to six sources at the receptor in the center, using the equation below.

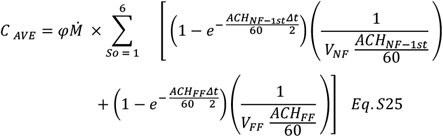

We can calculate the *ACH*_*NF*_ using the equation derived earlier for a hexagon:

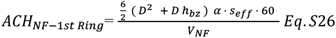

By substituting for the *V*_*NF*_,

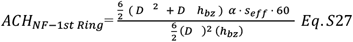

Which simplifies to:

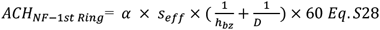

In the same way that a six source system was devised, a 12-source system that still keeps each person in the system *D* distance apart, but in this case is located two *D* away from the receptor. In this case, instead of a hexagon, a dodecagonal prism (12-sided prism) is drawn (**Fig. 3F**). In the 12-source system, we take 1/12th of the emissions and use 1/12 of the total dodecagonal prism volume. The *ACH*_*NF*_ is calculated using the dimension of the 1/12 triangular wedge which is derived as follows.

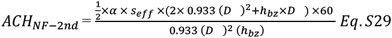

Which simplifies to:

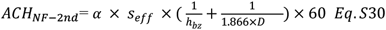

Successive rings, out to nine rings are included in CEAT, to allow up to a maximum of 270 people. Each ring adds 6 additional people more than the previous ring (i.e, the first ring holds 6 people, the second ring holds 12 people, the third ring holds 18, etc.) (**Fig S3E**). **Table S4** has the equations for the area of each of the triangular prisms, along with the equation used to calculate the *ACH*_*NF*_ for each ring.

Applying the superposition principle, the contribution of each person on the receptor at the center can be calculated. Going out to 60 sources (four rings) we get:

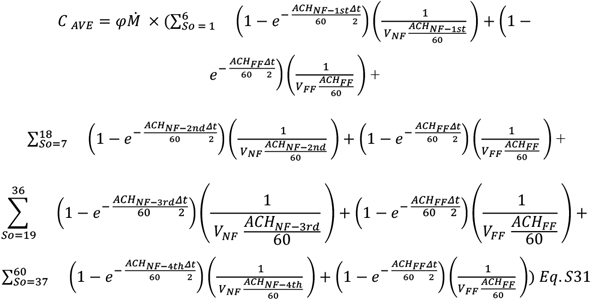

The formula may be simplified by pulling out the factors common in the two terms and rearranging as follows:

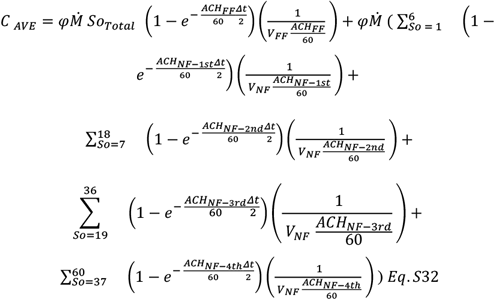

### Calculating the effective *ACH*_*FF*_ and *ACH*_*NF*_ to address sinks and turbulence

For the purposes of calculating the far-field concentration term, the *ACH*_*FF*_ should include any mechanisms that remove air from the space (e.g., natural ventilation, infiltration mechanical ventilation), mechanisms that remove the contaminant from the space (e.g., filtration and deposition), and mechanisms inactivate contaminants (e.g., reaction, temperature, humidity, radiation).

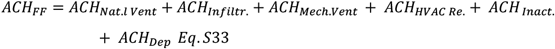

The *ACH*_*HVAC Re*._be based upon the flow rate and the portion of the recirculated air from which any contaminants has been removed (*ACH*_*HVAC Re*_ × *Ef*_*Filter*_):

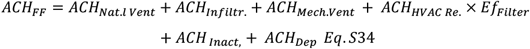

For the purposes of calculating the eddy diffusivity, the *ACH*_*FF*_ should only include mechanisms that result in actual air flow. So the *ACH*_*Inact*._ and *ACH*_*Dep*_ have not been included and the unreduced *ACH*_*HVAC Recirc*_ should be used, as shown.

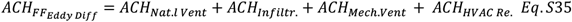

For the final *ACH*_*NF*_, the *ACH*_*Inact*._ and *ACH*_*Dep*_ should be added back in, as shown below for the 1st Ring of sources:

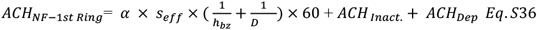

### Dose Model

As stated earlier, we employ a basic inhalation dose model:

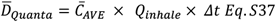

Where, 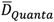 is the quantity of inhaled infectious material, 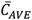 is average air concentration over the duration (mass/m^3^), *Q*_*inhale*_ the inhalation rate (m^3^/min), and *Δt* is the duration of exposure (min).

Since we are looking at this model from a worker safety perspective, we can also look at the total inhalation dose of all people in an activity space by multiplying the total number of people, assuming we are using, ideally, an average concentration and the same duration in the activity space.

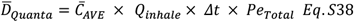

The concentration contributions are calculated for a person assumed to be at the center of a triangular grid where people are spaced equidistantly (based upon the distancing specified). We assume the concentration at the center is representative for all people in the group since: 1) each person’s location is likely not static during the activity and 2) exposure is driven mostly by the close-in sources (i.e., other people) and all people have close-in sources.

If we include mask effectiveness in the model, recognizing that there is an effect on both the inhalation side (1 − *Ef*_*in*_) and on the exhalation side (1 − *Ef*_*out*_), the equation takes the following form:

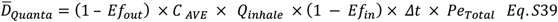

This equation calculates the total inhalation dose that a worst-case person (located at a receptor at the center of all rings) would receive if all people were emitting at a rate 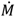 for the exposure duration. It assumes that all people are emitters (i.e., infected), when in fact only a few may be emitters. Based upon the *homogeneity property* of the *principle of superposition, φ*, in the expanded dose equation can be the likelihood that a person is infected, as shown below.

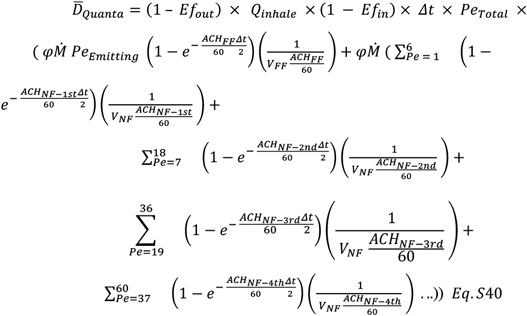

or written more succinctly,

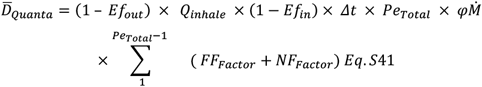

where,

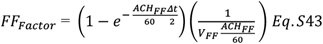

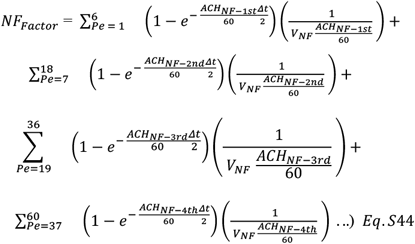

Additional terms can be added to Eq. S44 for each hexagonal ring as more people are added. CEAT allows up to 250 people.

### Impact of Prevalence of Infection in Community

Critical to the exposure assessment is the consideration of the likelihood that any individual member of the group is infectious at the start of the scenario or modeled event, with the likelihood of infection represented by the variable *φ*. In the CEAT model, the range of likelihood of infectiousness in the group can range from 1.0 (certain infectiousness) on the high end, to a value on the low end that is 100 times less than what is estimated as the community average infectiousness. In all of the cases, we assume that at least one person is not infectious, so the population that could be infectious is the size of the group, Pe, minus 1.

We estimate the community average infectiousness by using the reported 7-day average per 100,000 of diagnosed cases (*Cases*_*Per* 100000_), an estimate of the ratio of the undiagnosed cases over the diagnosed cases (*R*_*Undiag*_), and the average length of an infectiousness in days, (*D*_*Inf*_), multiplied by the subgroup factor, which is the adjustment of the subgroup’s rate of infectiousness as compared to the rate of infectiousness of the community.

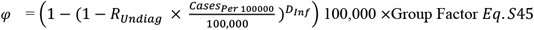

We assume that within a community, the population can be subdivided into subpopulations as follows:

1. Group Factor = 0.01 The Group is composed of people who, prior to the event are estimated as having a likelihood COVID-19 infection that is 100 times lower than the community’s average due to their adhering to public health guidance on distancing, masking, and exposure to crowds/people.
2. Group Factor = 0.1 The Group is composed of people who, prior to the event are estimated as having a likelihood COVID-19 infection that is 10 times lower than the community’s average due to their adhering to public health guidance on distancing, masking, and exposure to crowds/people.
3. Group Factor = 1 The Group is composed of people who, prior to the event are estimated as having a likelihood COVID-19 infection that is equal to the community’s average.
4. Group Factor = 0.1 The Group is composed of people who, prior to the event are estimated as having a likelihood COVID-19 infection that is 10 times higher than the community’s average due to their not adhering to public health guidance on distancing, masking, and exposure to crowds/people.
5. *φ* = 1 The group is composed of people who are known to be infectious.

### Impact of Variants

We handle the current community prevalence of variants and the relative infectiousness of the prevalent variants by assuming that some variants may be significantly more or less transmissive than other variants. For the fraction of total cases of more infectious variants, we can adjust the fractional exposure upward or downward to account for its infectiousness.

### Efficacy of Immunity

Immunities, including vaccination and recovered cases, are addressed in two ways:

1. It reduces the rate of virus shedding of immunized persons who do become infected, thus reducing the emission rate, 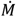, for the fraction of people with immunity; this is based upon a 3 times reduction in shedding observed by [85].
2. The immunity is treated as a barrier to infection with an effectiveness that is equal to its published efficacy based conceptually on the model used by the EPA for dose and exposure definition [29].

We are assuming that immunity gained by recovery from COVID is equal to the immunity gained from vaccination.

### Efficacy of Testing

We address the efficacy and timing of testing regimes, relative to the days an individual is expected to be infectious. We assume that if an individual is infectious, at the time of the event, the timing of the infection prior to the event is a uniform distribution. For example, if *D*_*Inf*_ = 5, and they were tested three days prior to the event, there is a 3/5 chance they were infected when they were tested and a 2/5 chance they got infected after they were tested (in the two subsequent days before the event). Assuming a testing false negative rate (*R*_*False Neg*._) of 10%, the testing adjustment factor, which assumes testing was performed three days before the event, is computed as follows:

- If *D*_*Inf*_ < 3, there is no adjustment to the likelihood that an individual is infectious because testing was performed prior to anyone becoming infectious.
- If the *D*_*Inf*_ ≥ 3, the testing adjustment is computed as the weighted likelihood of either (a) having been infected at the time of testing *and* obtaining a false negative test or (b) becoming infected after the test:

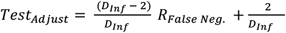

where *R*_*False Neg*._is currently set to 0.10.

### Relative Dose Ratio Approach: How we establish the baseline

Rather than directly calculating a dose-response, we use a comparative dose approach. We compare all scenarios to a baseline scenario discussed in **Table S2**. The model’s results are aligned with the US OSHA classifications of exposure risks [34], by benchmarking the dose calculations to a baseline scenario that is considered high risk by US OSHA. We define the baseline scenario to represent a person (i.e., medical worker) who is exposed to a COVID-19 infected person. We apply assumptions to this scenario, addressing each of the factors in **Table S2**, to arrive at a baseline inhalation dose value. The inhalation dose for other scenarios is compared to the baseline dose by a simple ratio. Below is the full ratio equation with the “*i th”* scenario in the numerator and the baseline (BL) in the denominator. We can rearrange the terms in each of the *i* scenario 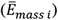 and the baseline 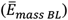:

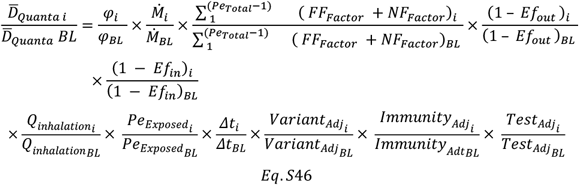

### Emission Rate Approach

Deterministic dose-response models provide estimations of the intake dose and estimations of the probability of infection for the intake dose. These models require a means of quantifying the dose and quantifying the pathogen-host interaction via a dose response (i.e., a *tolerance dose* – the dose above which someone is certain to be infected or a *threshold dose* – minimum dose needed to initiate a chance of infection in any person) [15]. To calculate risks using a dose-response approach, similar to what was done by Parhizkar, et al., 2021, the following is needed: (1) an explicit mass rate or particle count rate emitted from an infected person, (2) information on particle size emitted and particle size distribution, and (3) the explicit response threshold dose or tolerance dose. Determining these data requires environmental measurement and epidemiological studies of transmission. While CEAT is also based upon a deterministic dose-response framework, it does not use explicit values for emission rate and dose response. Instead it calculates a dose ratio (using Equation S44), based upon comparing a baseline scenario that has been defined as high risk to an evaluated scenario (i.e., i^th^ scenario). This simplification provides a means to rapidly deploy a comprehensive risk model during an infectious disease outbreak ahead of public health and medical authorities having detailed data on the explicit viral emission rates and dose responses. The CEAT model does, however, require that a public health authority (e.g., Occupational Safety and Health Administration (OSHA), Centers of Disease Control and Prevention (CDC), or other governmental health department) or other expert defines an exposure and dose scenario that is consistent with high risk exposure. In CEAT, we have used the OSHA classifications of exposure risks [34] for this purpose.

While the ratio model does not directly use Wells-Riley approach, it does benefit from the data that have been empirically-derived from use of the Wells-Riley approach, allowing us to adjust the CEAT dose ratio results and exposure risk results for various activities and vocalization intensities. We use the back-calculated quanta per hour from Buonanno, et al., 2020a and Buonanno, et al., 2020b to inform the ratio of emission rates, 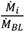.We make the assumption that these empirically-derived ratios would be correlated with explicit mass or particle count ratios that would be appropriate for deterministic dose-response models.

It is instructive to note that the CEAT approach does not require a means of varying the emission rate ratios. If Wells-Riley-derived emissions for various activities and vocalization intensities were not available, the assumption could be made that emission rate was constant (i.e., 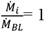). All of the other ratio factors in Eq 46 could still be used to evaluate the i^th^ dose scenario versus the baseline scenario. The majority cases that the CEAT was employed assumed that the Step 5 vocalization intensity was “standing and speaking” which uses in 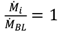 in the model’s calculations. The fact that Wells-Riley-derived data are not essential to use CEAT is a benefit of the CEAT approach.

## QUANTIFICATION AND STATISTICAL ANALYSIS

### Gathering Scenario

To determine the gathering scenario, we considered three different counties for the date of 1/31/2022 that were in different regions of the US with very low COVID-19 cases (Montgomery County, MD), very high COVID-19 cases (Knox County, TN), and a county with cases in between the two (Suffolk County, MA). In this analysis, we estimated that a typical gathering will last 5 hours and can be held both indoors or outdoors. The indoor scenario is considered to take place in a room (i.e., 30ft x 30ft x 9ft or 9.14m x 9.14m x 2.74m). We utilized the COVID ActNow tracker [86] to determine the latest number of cases and vaccination rates on 1/31/2022. In addition, we utilized CDC’s Nationwide Commercial Laboratory Seroprevalence Survey [86] to determine the current population recovered from COVID-19, and CDC’s Variant Proportions Tracker [86] to determine the estimated percentage of existing SARS-CoV-2 variants that exist in the infected population in each region. At the time of analysis for all counties the Omicron variant accounts for >99% of COVID-19 cases [86]. It is estimated that the Omicron variant is 440% more transmissible than the original SARS-CoV-2 reference strain [87]. We analyzed for the following different parameters to account for multiple different scenarios that gatherings can take place: distancing ranging from 1.5ft to 10ft, masks usage (i.e., no masks, average masks, and N95/KN95 masks), and if the group of people are either “following all public health guidance” or “equal to the community average”. In addition, we also considered testing to be included with one of the scenarios. All data was recorded in Microsoft Excel 2019 and all data analyses were completed using R version 4.0.3, RStudio version 1.4.1717, and ggplot2 v3.3.5 [88].

### NASA Ames Research Center CEAT Tool Usage

In initial assessments, utilizing CEAT V B.6 that was released on November 25, 2020 Step 1 (“The group is composed of people who…”) was generally selected as “You think are following all public health guidance”. Step 2 (“Number of People Sharing Activity Space”) was set to the requested number of personnel required to conduct the operation in-person. In general this was 2 to 4 people per location per operation. Selected distance (Step 3) was set to 6 feet (“-6 distancing adjustment”) unless specified otherwise. For Mask Efficacy (Step 4) “cloth masks” worn by all personnel were selected as cloth was the most likely utilized (−5 and −3, respectively). Very few projects were using surgical masks and masks were required to be worn by everyone on campus at this time. Vocalization (Step 5) and breathing (Step 6) adjustment rates, as well as Duration of Activity (Step 7) were based on the operations reported in the RTOW plan submission. Typical operations are conducted while “standing”, “speaking”, and “passive” (0 and 0) for 8 hours. Ventilation rates (Step 8) and Adjustment for room sizes (Step 9) were based on location of the operation reported in the RTOW plan submission. Step 10 (“Calculate Adjustment to Local Community’s Current Conditions”) was based on the State of California [46]. The California case rate was chosen instead of the local county case rate as the majority of the NASA ARC workforce resides in the general Bay Area which encompasses nine counties, some of which have weekly case rates more similar to California than to the local county. After inputting these desired values for the variables in Steps 1-10, the relative exposure ratio for a given condition was recorded and analyzed in Microsoft Excel 365.

CEAT V B.14 was released on December 13, 2020 the inputs were similar to that of V B.6, the difference being that for Step 9 actual room dimensions could be entered.

CEAT V B.29 was released on May 6, 2021 Step 1 (“The group is composed of people who…”) was selected as “Are following all public health guidance”. However, since the percent vaccination rate was unknown, it was not checked. Step 2 (“Number of People Sharing Activity Space”) was set to the requested number of personnel required to conduct the operation in-person. In general this was 2 to 4 people per location per operation. Selected distance (Step 3) was set to 6 feet unless specified otherwise. For “Mask Type and Prevalence” (Step 4) “cloth masks” worn by all personnel were selected as cloth was the most likely utilized. Very few projects were using surgical masks and masks were required to be worn by everyone on campus for all but a 6 week window where masks were optional for vaccinated personnel. Vocalization (Step 5) and breathing (Step 6) adjustment rates, as well as Duration of Activity (Step 7) were based on the operations reported in the RTOW plan submission. Typical operations are conducted while “standing”, “speaking”, and “passive” for 8 hours. Ventilation rates (Step 8) and Adjustment for room sizes (Step 9) were based on location of the operation reported in the RTOW plan submission. Step 10 (“Calculate Adjustment to Local Community’s Current Conditions”) was based on the State of California [46,89] and variant information was input from CDC data [90]. When variant prevalence was introduced into the CEAT in later iterations, the three most prevalent variants in Health and Human Services (HHS) Region 9 were used [90]. Specifically, the variant prevalence data from Nowcast was utilized. Instead of utilizing the predetermined variants provided in the CEAT, NASA ARC input data from the three most prevalent variants in the HHS Region 9. The “Protection Effectiveness of Immunity (%)” in Step 10 was set to 66% based on published research regarding the Pfizer-BioNTech, Moderna, and Janssen vaccine against the Delta variant [91]. After inputting these desired values for the variables in Steps 1-10, the relative exposure ratio for a given condition was recorded and analyzed in Microsoft Excel 365. Although CEAT V B.32 was released on August 29, 2021 it was not used in this analysis. To generate a graphical representation of the data (**Fig. 4**) we associated numerical values to the different parameters in the table and utilized R version 4.03, RStudio version 1.4.1717 with the following R packages: ggplot2 v3.3.5 [88].

For the longitudinal review of the NASA ARC “Centerwide Accepted Median Exposure Risk Ratio” in relation to the community case rates CEAT V B.6, V B.14, and V B.29 were utilized, this was dependent on the newest version available. Initial inputs at the time of the RTOW plan were utilized and Step 10 (“Calculate Adjustment to Local Community’s Current Conditions”) rates were updated on a biweekly basis based on the State of California [46]. The median of all project exposure risk ratios was used instead of the average to account for the high fluctuations in exposure risk ratios. Hypothetical exposure risk ratios were back-calculated to March 2020. Only projects that had been approved to RTOW, along with projects that were deemed mission essential and were exempt from the work from home policy (e.g., Security Guards, Security Operations Center) were included in the calculated biweekly median risk ratio. The relative exposure ratio for a given condition was recorded, the median exposure ratio was calculated biweekly, and the correlation coefficient compared to the community case rates was calculated in Microsoft Excel 365. The median of all project exposure risk ratios was used instead of the average to account for the high fluctuations. Although the CEAT was not used at NASA ARC until December 2020, hypothetical exposure risk ratios were back-calculated to March 2020, when NASA ARC enacted their mandatory work from home policy, for each project using the known historic California case rates. Only projects that had been approved to RTOW, along with projects that were deemed mission essential and were exempt from the work from home policy (e.g., Security Guards, Security Operations Center) were included in the calculated biweekly median exposure risk ratio. A plot was generated for this data (**Fig. 5**) using R version 4.0.3, RStudio version 1.4.1717 with the following R packages: ggplot2 v3.3.5 [88].

---

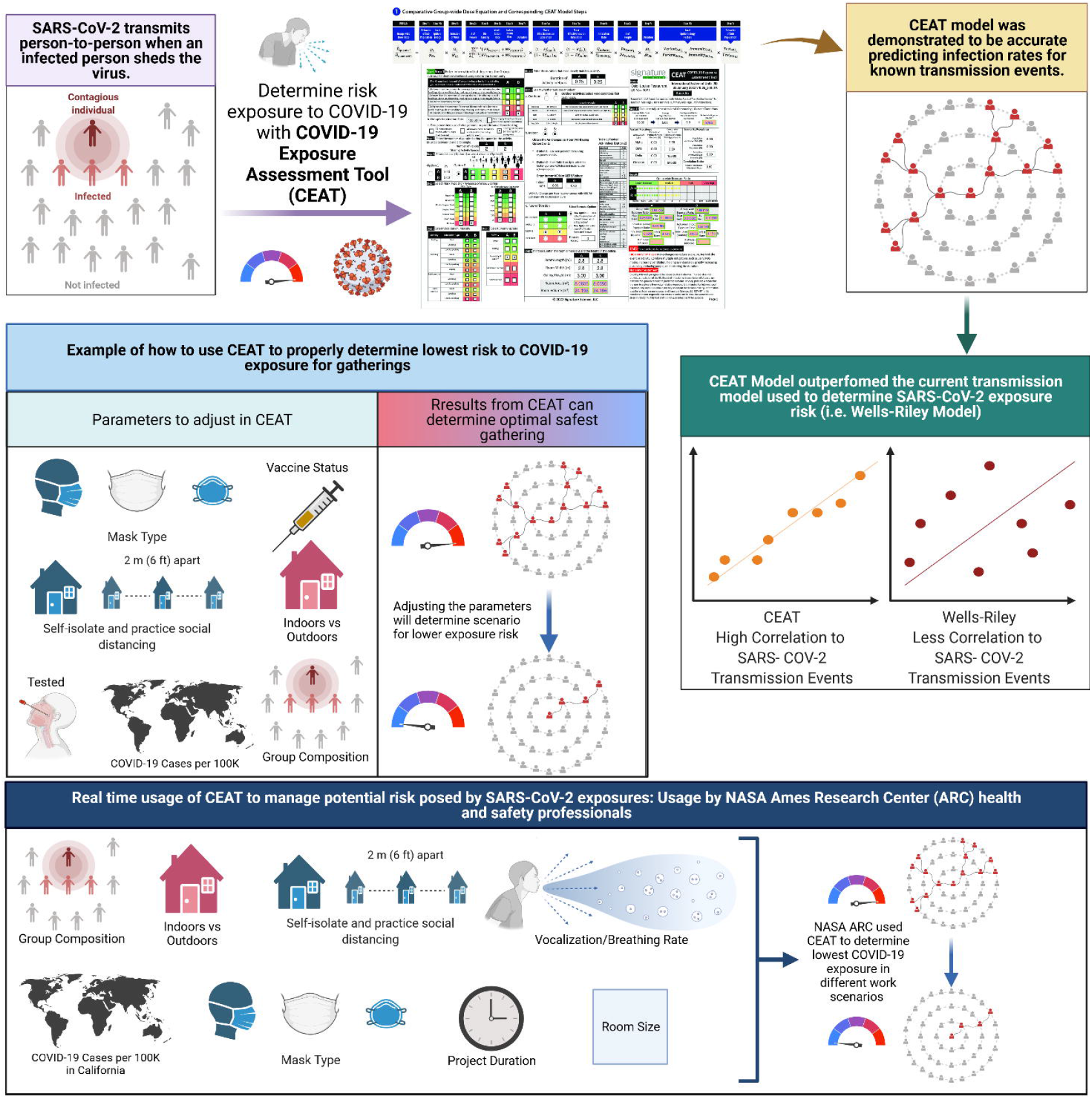

