## Supplemental Figures and Tables for "Covid-19 Exposure Assessment Tool (CEAT): Easy-to-use tool to quantify exposure based on airflow, group behavior, and infection prevalence in the community"

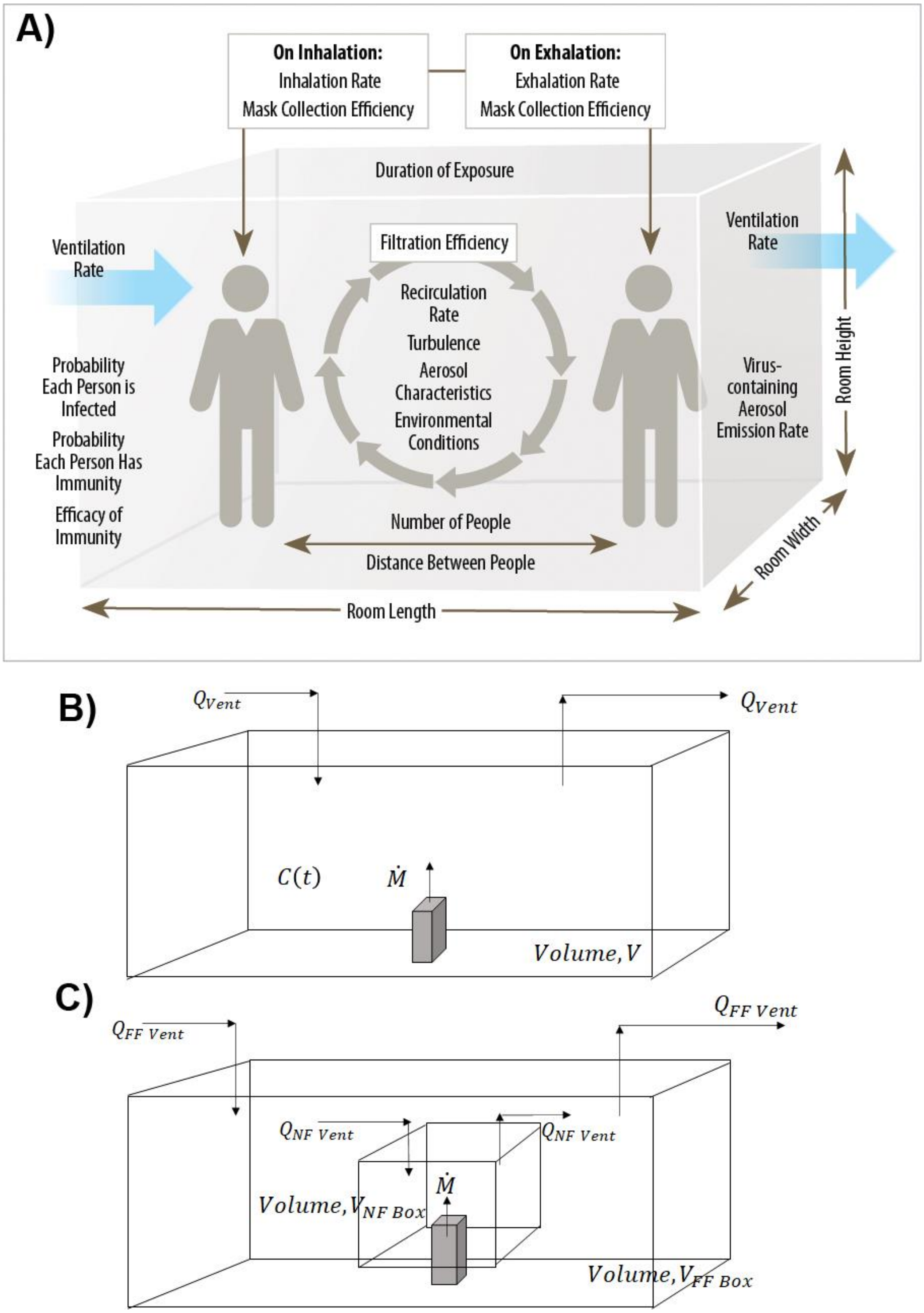


**Figure S1. The different box models for CEAT. A)** Factors Included in COVID-19 Exposure Assessment Tool Interface (CEAT). A summary of the factors and mechanisms affecting the comparative dose and exposure risks. **B)** Single Zone Well-Mixed Box Model. Basic box model assumes emissions are instantaneously well mixed. **C)** Near Field (NF) and Far Field (FF) Box Model. "Box-within-a-box" approach provides localized higher concentration in the vicinity of the source.

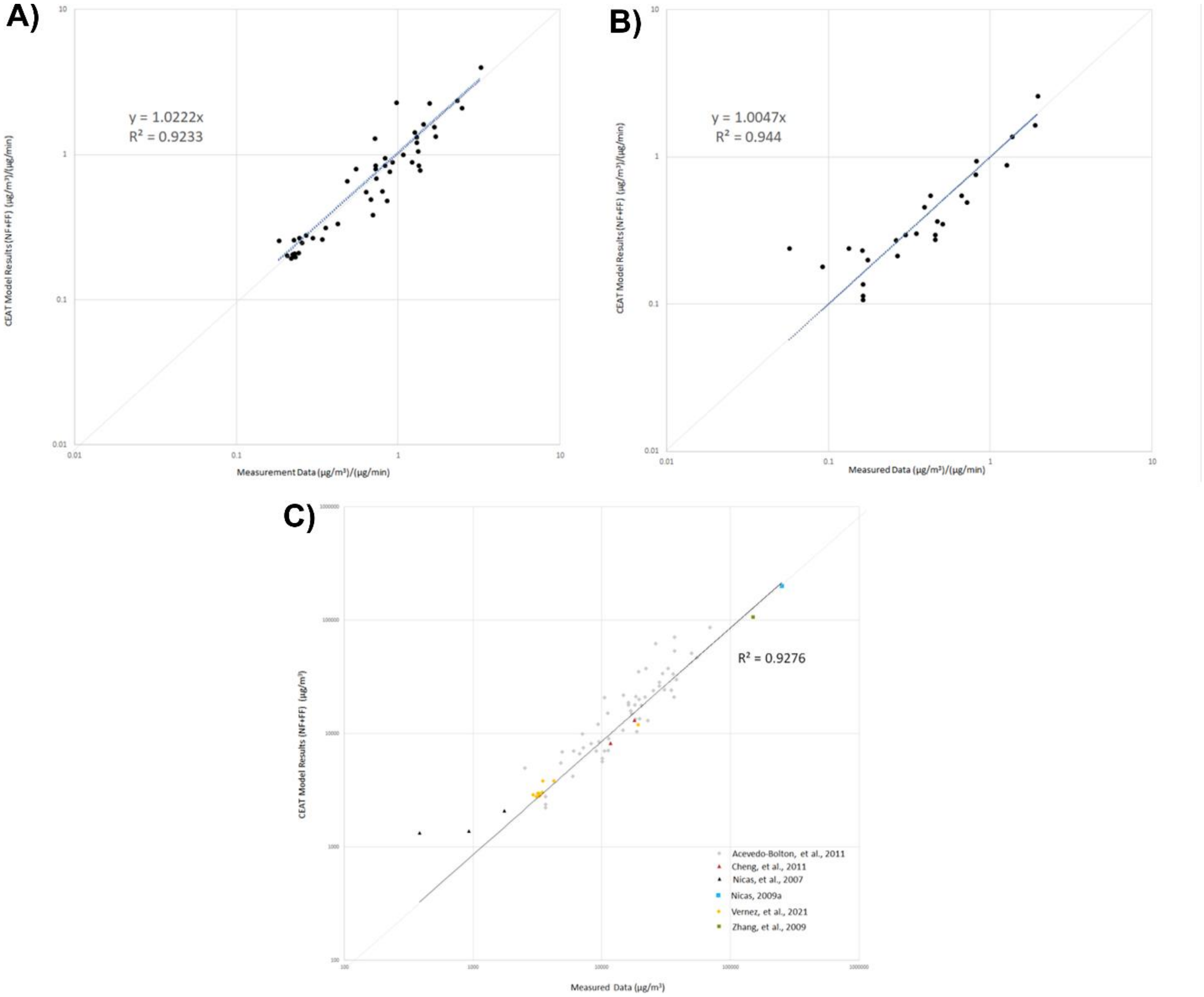


**Figure S2. CEAT Concentration Model Performance. A)** Modeled results vs. measured concentrations using CEAT concentration model, applying the relationship between ACH, room size and eddy diffusivity according to Venkatram and Weil, 2021 in cases when ACH was less than or equal to than 0.75 $hr^{-1}$**.** Results are normalized by dividing by the emission rate. **B)** Modeled results vs. measured concentrations using CEAT concentration model, applying the relationship between ACH, room size and eddy diffusivity according to Foats et al., 2021 in cases when ACH was greater than 0.75 $hr^{-1}$ and assuming 4 vents per 100 $meters^2$**.** Results are normalized by dividing by the emission rate. **C)** Modeled results vs. measured concentrations using CEAT concentration model for all cases.

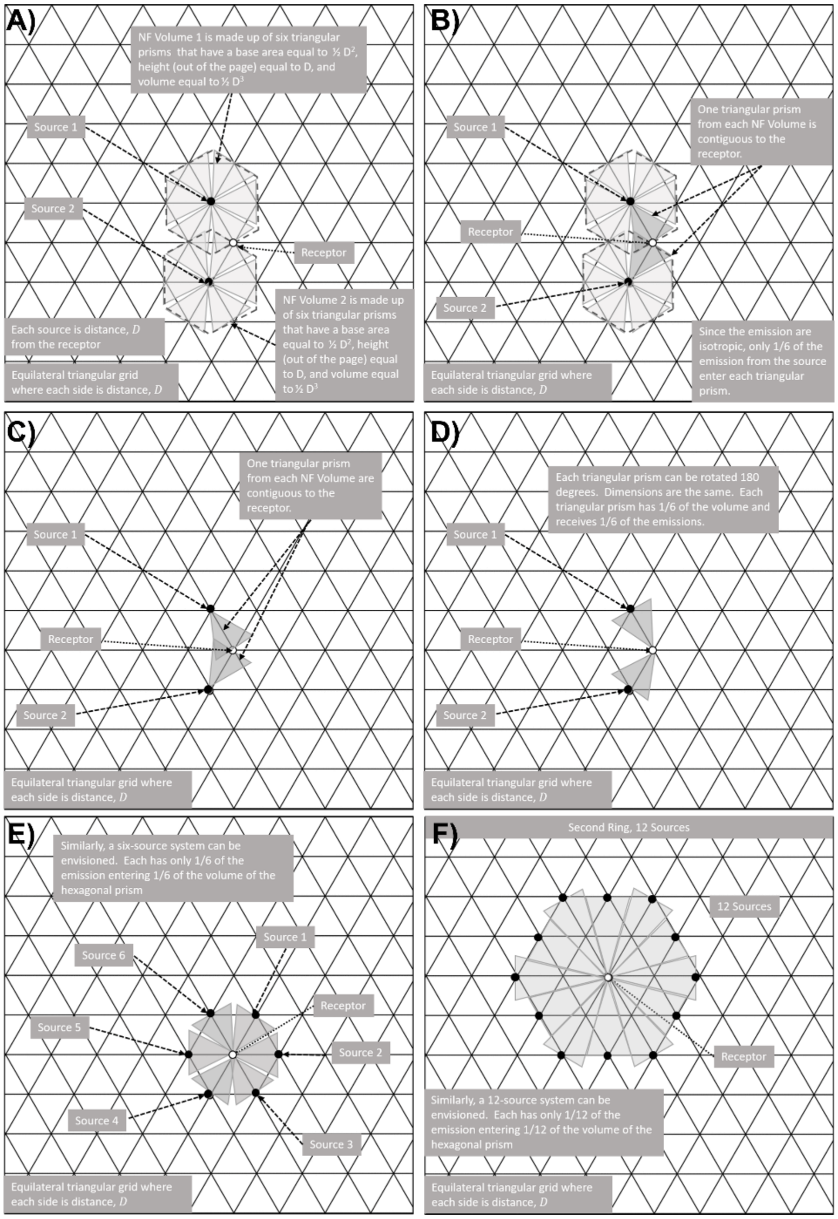


**Figure S3. Dimensions and sources for the box model. A)** Near Field (NF) and Far Field (FF) Box Dimensions with Two People. **B)** The application of the principle of superposition with Near Field (NF) and Far Field (FF). **C)** Near Field (NF) Triangular Prisms for the 1st Ring of the Group. NF triangular prisms for the 1st ring of the group that have a height and base of $D_{tot}$, in a triangular grid with each side length of $D_{tot}$. The first ring's triangular prisms have a base area of A1 = ½ $D_{tot}{}^2$. **D)** Near Field (NF) Triangular Prisms for the 2nd Ring of the Group. NF triangular prisms for the 2nd ring of the group that is equally spaced $D_{tot}$. The second ring's triangular prisms have areas A2 = ½ $2D_{tot}{}^2$ and A2 = ½ $\sqrt{3}\, D_{tot}{}^2$. These average to $A2 = 0.9330\, D_{tot}{}^2$. **E)** Source perspective with two sources shown. Under an assumption of isotropy assuming no predominate flow and a sufficient averaging period, the sources emit in all directions equally. **F)** The same two triangles that impact the receptor in the source perspective can be turned 180 degrees and are part of the potential set of triangles in each ring. The dimensions and parameters are identical between both views.

**Table S1. OSHA Risk Classifications.** OSHA's classifications offer a means of comparing exposures to a scenario that can be defined as high risk (US OSHA 2020).

| | Lower Risk | Medium | High | Very High |
|---|---|---|---|---|
| OSHA Category (OSHA, March 2020) | Lower exposure risk (caution) jobs are those that do not require contact with people known to be, or suspected of being, infected with SARS-CoV-2 nor frequent close contact with (i.e., within 6 feet of) the general public. Workers in this category have minimal occupational contact with the public and other coworkers. | Medium exposure risk jobs include those that require frequent and/or close contact with (i.e., within 6 feet of) people who may be infected with SARS-CoV-2, but who are not known or suspected COVID-19 patients. In areas where there is ongoing community transmission, workers in this category may have contact with the general public (e.g., schools, high-population-density work environments, some high-volume retail settings). | High Exposure Risk High exposure risk jobs are those with high potential for exposure to known or suspected sources of COVID-19. Workers in this category include: ■ Healthcare delivery and support staff (e.g., staff who must enter patients' rooms) exposed to known or suspected COVID-19 patients. ■ Medical transport workers moving known or suspected COVID-19 patients in enclosed vehicles. ■ Mortuary workers involved in preparing the bodies of people who are known to have, or suspected of having, COVID-19. | Very High Exposure Risk Very high exposure risk jobs are those with high potential for exposure to known or suspected sources of COVID-19 during specific medical, postmortem, or laboratory procedures. Workers in this category include: ■ Healthcare workers performing aerosol-generating procedures on known or suspected COVID-19 patients. ■ Healthcare or laboratory personnel collecting or handling specimens from known or suspected COVID-19 patients. ■ Morgue workers performing autopsies, which generally involve aerosol-generating procedures |
| | Lower Exposure | Medium | High | Very High |
| Corresponding CEAT Category | Based upon the US community exposure of approximately 4 cases per 100,000 which occurred in late March 2020, CEAT would calculate a low exposure dose ratio for unprotected community members that behaved consistent with this OHSA category. | Based upon the US community exposure of approximately 4 cases per 100,000 which occurred in late March 2020, CEAT would calculate a medium exposure dose ratio for unprotected community members that behaved consistent with this OHSA category. | CEAT would calculate a high exposure for unprotected people in close contact with known or suspected COVID-19 patients. | CEAT would calculate a high exposure for unprotected people in close contact with known or suspected COVID-19 patients that were in high aerosol-generating activities (heavy exertion, speaking loudly) in close proximity. |

**Table S2. Baseline Scenario Approach.** Mechanisms affecting the exposure risk.

| Factors considered in the Baseline Exposure Dose Calculation | CEAT Step # | Value and Basis assumed for the Baseline Scenario |
|---|---|---|
| Likelihood of Infectious persons present in the group | 1 | 100% that 1 known infection is assumed in the room. |
| Number of people in the group | 2 | 2 people. |
| Distance between people | 3 | ~1 m ((~3 ft) based on estimated average distance during the event that is less than 6 feet |
| Mask effectiveness | 4 | No masks are worn |
| Mask compliance on the group | 4 | No masks worn |
| Emission rate of Infectious aerosols released through respiration | 5 | 11.4 quanta per hour, associated with speaking while standing. (Buonanno, et al., August 2020) (Buonanno et al., December 2020) |
| Inhalation rate | 6 | 13 liters per min (0.46 cfm) based on assumption of light intensity (US EPA, 2015) |
| Duration of exposure | 7 | 15 minutes, corresponding to the time defined by CDC contact tracing as a significant exposure (CDC, February 2020) |
| Indoors or outdoors activity | 8 | Indoor |
| Ventilation rates (air changes per hour [ACH] or air exchange rate [AER]) | 8 | 6 ACH based upon CDC guidance for health care facilities. (CDC, 2019) |
| Aerosol settling rate | 8 | Add 0.24 to the ACH for the removal by deposition on surfaces (CIRES, 2020) |
| Virus degradation rate | 8 | Add 0.63 to the ACH for the viral aerosol degradation assuming 20.1 degrees C and 38% humidity (CIRES, 2020) |
| Recirculating room filtration rate and removal efficiency | 8 | No additional contribution to ACH from recirculating air filtration is assumed. |
| Volume of room or activity space | 9 | Room size assumed to be a small examination room that was 9 square meters (96.8 square feet) with a ceiling height of 2.74 meter (9 feet) |
| Prevalence of COVID-19 in the community | 10 | Since the likelihood of Infectious persons present in the group is 100%, the prevalence of COVID-19 in the community is not used in the model |
| Difference in the variants transmission rates versus wild type virus | 10 | The wild type virus is assumed. so no adjustment is made. |
| Impact of community's or group's immunity from recovery and vaccination | 1 and 10 | No immunity is assumed, so no adjustment is made. |
| Impact of surveillance testing for the group | 1 | No surveillance testing is assumed, so no adjustment is made |

### Table S4. Comparison of Three Simple Approaches to Modeling a Continuous Point Release

| Gaussian solution: Source above a surface with a reflection source | NF/FF with a Spherical NF Volume | NF/FF with a Hexagonal Prism NF Volume |
|---|---|---|
| Starting with the Gaussian equation for a continuous point release suspended above a surface at a height of H when there is no advection (Stockie, 2011).<br>$C\ (x, y, z)\ = \frac{\dot{M}}{4\pi\ K}\left(\frac{1}{\sqrt{x^2\ +\ y^2\ + (z-H)^2}} + \frac{1}{\sqrt{x^2\ +\ y^2\ + (z+H)^2}}\right)$<br>where x,y, and z are distances on a cartesian coordinate system. If we then set y=0 and z=H, we have an equation to predict the concentration along the x-axis as x increases to x = D, the results converges on for large values of D.<br>$C(x)\ = \frac{\dot{M}}{2\pi\ K\ D}$<br>This derivation is considered valid when K is constant in the system<br>$C(x)\ = \frac{\dot{M}}{2\pi\ K\ D}$<br>Result:<br>$C(x)\ = \frac{\dot{M}}{2\pi\ K\ D}$ | Apply the NF/FF model where the size of the space is large as compared to the NF Volume (i.e., $V_{FF}\ >> V_{nF}$ ) (Nicas, 2009a), starting with the original form of the NN/FF model at equilibrium<br>$C_{eq}\ = \frac{\dot{M}}{\beta} + \frac{\dot{M}}{Q_{FF}}$<br>and assuming a space is very large, we can assume that $\frac{\dot{M}}{Q_{FF}} >> \frac{M}{Q_{FF}}$, such that<br>$C_{eq}\ = \frac{\dot{M}}{Q_{NF}}$<br>If we apply the FSA method:<br>$C_{eq}\ = \frac{\dot{M}}{\frac{1}{2}\ FSA\ \times\ S}$<br>and then assume the Near Field volume is a sphere with a surface area, A = $4\pi R^2$ a distance, D, we get<br>$C_{eq}\ = \frac{M}{2\pi\ D^2\ s} = \frac{\dot{M}}{2\pi\ (D\ \times\ s)\ D}$<br>Result:<br>$C_{eq}\ = \frac{\dot{M}}{2\pi\ K\ D}$ if<br>$K\ =\ D\ \cdot\ s$ | Apply the NF/FF model where the size of the space is large as compared to the NF Volume (i.e., $V_{FF}\ >> V_{nF}$ ) (Nicas, 2009a), starting with the original form of the NN/FF model at equilibrium<br>$C_{eq}\ = \frac{\dot{M}}{Q_{NF}} + \frac{\dot{M}}{Q_{FF}}$<br>and assuming a space is very large, we can assume that $\frac{\dot{M}}{Q_{FF}} >> \frac{M}{Q_{FF}}$, such that<br>$C_{eq}\ = \frac{\dot{M}}{Q_{NF}}$<br>If we apply the FSA method:<br>$C_{eq}\ = \frac{\dot{M}}{\frac{1}{2}\ FSA\ \times\ S}$<br>and then assume the Near Field volume is a Hexagon with a surface area, A = [6 × (2 × ½ $D^2$)+ (D $h_{bz}$)] a distance, x, we get<br>$C_{eq}\ = \frac{\dot{M}}{\frac{1}{2}\ [6\ \times\ (2\ \times\ ½\ D^2\ ) + (□\ h_{bz})]\ \times\ S}$<br>$= \frac{\dot{M}}{\frac{1}{2}\ 6\ (D^2\ +\ D\ h_{bz}\ )\ \times\ S}$<br>If $D\ = h_{bz}$ , then the equation is<br>$C_{eq}\ = \frac{\dot{M}}{6\ \ D \times D \times S}$ or<br>Result:<br>$C_{eq}\ = \frac{\dot{M}}{6\ \ K\ D}$ if<br>$K\ =\ D\ \cdot\ s$ and note that $6 \cong 2\pi$ |

**Table S5. Triangular Prism Parameters and Equations for Each Ring**

| Ring # | People per Ring | Cumulative People | Triangles Per Ring | Cumulative Triangles | Triangles per People by Ring | Equation for the Area of NF Triangular Prism | Equation for Area of NF Triangular Prism in Terms of A1 | ACH NF Equation for each Ring |
|---|---|---|---|---|---|---|---|---|
| **1** | 6 | 6 | 6 | 6 | 1 | A1 =0.5 $(D_{tot})^2$ | A1 = A1 | $\alpha \times s \times (\frac{1}{h_{bz}} + \frac{1}{D_{tot}}) \times 60$ |
| **2** | 12 | 18 | 18 | 24 | 3/2 | A2 = 0.933 $(D_{tot})^2$ | A2 = A1 x 1.866 | $\alpha \times s \times (\frac{1}{h_{bz}} + \frac{1}{1.866 \times D_{tot}}) \times 60$ |
| **3** | 18 | 36 | 30 | 54 | 5/3 | A3= 1.40$(D_{tot})^2$ | A3 = A1 x 2.799 | $\alpha \times s \times (\frac{1}{h_{bz}} + \frac{1}{2.799 \times D_{tot}}) \times 60$ |
| **4** | 24 | 60 | 42 | 96 | 7/4 | A4= 1.87 $D_{tot}{}^2$ | A4 = A1 x 3.732 | $\alpha \times s \times (\frac{1}{h_{bz}} + \frac{1}{3.732 \times D_{tot}}) \times 60$ |
| **5** | 30 | 90 | 54 | 150 | 9/5 | A5 = 2.33 $D_{tot}{}^2$ | A5 = A1 x 4.665 | $\alpha \times s \times (\frac{1}{h_{bz}} + \frac{1}{4.665 \times D_{tot}}) \times 60$ |
| **6** | 36 | 126 | 66 | 216 | 11/6 | A6 = 2.80 $D_{tot}{}^2$ | A6 = A1 x 5.598 | $\alpha \times s \times (\frac{1}{h_{bz}} + \frac{1}{5.598 \times D_{tot}}) \times 60$ |
| **7** | 42 | 168 | 78 | 294 | 13/7 | A7 = 3.27 $D_{tot}{}^2$ | A7 = A1 x 6.531 | $\alpha \times s \times (\frac{1}{h_{bz}} + \frac{1}{6.531 \times D_{tot}}) \times 60$ |
| **8** | 48 | 216 | 90 | 384 | 15/8 | A8 = 3.73 $D_{tot}{}^2$ | A8 = A1 x 7.464 | $\alpha \times s \times (\frac{1}{h_{bz}} + \frac{1}{7.464 \times D_{tot}}) \times 60$ |
| **9** | 54 | 270 | 102 | 486 | 17/9 | A9 = 4.20 $D_{tot}{}^2$ | A9 = A1 x 8.397 | $\alpha \times s \times (\frac{1}{h_{bz}} + \frac{1}{8.397 \times D_{tot}}) \times 60$ |
